# Returning Actionable Genomic Results in a Research Biobank: Analytic Validity, Clinical Implementation and Resource Utilization

**DOI:** 10.1101/2021.08.03.21261489

**Authors:** Carrie L. Blout Zawatsky, Nidhi Shah, Kalotina Machini, Emma Perez, Kurt D. Christensen, Hana Zouk, Marcie Steeves, Christopher Koch, Melissa Uveges, Janelle Shea, Nina Gold, Joel Krier, Natalie Boutin, Lisa Mahanta, Heidi L. Rehm, Scott T. Weiss, Elizabeth W. Karlson, Jordan W. Smoller, Matthew S. Lebo, Robert C. Green

## Abstract

Over 100 million research participants around the world have had research array-based genotyping (GT) or sequencing (GS), but only a small fraction of these have been offered return of actionable genomic findings (gRoR). Between 2017 and 2021, we analyzed genomic results from 36,417 participants in the Mass General Brigham Biobank and offered to confirm and return pathogenic and likely pathogenic variants (PLPVs) in 59 genes. Variant verification prior to patient recontact revealed that GT falsely identified PLPVs in 44.9% of samples, and GT failed to identify 72.0% of PLPVs detected in a subset of samples that were also sequenced. GT and GS detected verified PLPVs in 1% and 2.5% of the cohort, respectively. Of 256 participants who were alerted that they carried actionable PLPVs, 37.5% actively or passively declined further disclosure. 76.3% of those carrying PLPVs were unaware that they were carrying the variant and over half of those met published professional criteria for genetic testing but had never been tested. This gRoR protocol cost approximately $129,000 USD per year in laboratory testing and research staff support, representing $14 per participant whose DNA was analyzed or $3,224 per participant in whom a PLPV was confirmed and disclosed. These data provide logistical details around gRoR that could help other investigators planning to return genomic results.

## INTRODUCTION

Research biobanks and other human research studies that collect and analyze DNA are increasingly confronted with the question of whether and how to return actionable genomic results to individual participants (gRoR). A majority of research participants^1-3^ and researchers^4; 5^ favor returning such results to participants, and many research studies that collect genomic data have written policies encouraging the return of actionable genomic results to participants (gRoR).^6-9^ Yet the vast majority of such studies in the US and around the world have not implemented gRoR due to uncertainties around how to consent participants, which genes to select for return, how to analyze, classify and report research variants, the logistics of recontacting participants, regulatory requirements necessitating the confirmation of research results, the transition of research participants into an appropriate clinical workstream and the effort and cost associated with each of these steps.^10-17^ Despite these challenges, it is likely that research participants will increasingly expect gRoR in genomic research.^18; 19^ For example, the NIH sponsored *All of Us* Research Program has announced that it will sequence and return actionable genomic results to 1 million Americans,^20^ adopting a process similar to that described in this article, and a 2018 National Academies of Science, Engineering and Medicine report predicted “the return of research results will soon become an integral part of the research enterprise” and stressed the need for detailed descriptions of consent practices, technical standards, participant preferences and resourcing for returning research results.^21^

Research studies and biobanks that have elected to return genomic information to research participants typically share common themes and workflows.^22-25^ First, participants must explicitly accept or decline gRoR at enrollment, or if this choice was not presented at enrollment, they must later be re-consented for gRoR. Next, a list of genes associated with actionable hereditary conditions is selected for analysis and potential return. Then, genotyping or sequencing data are filtered and interpreted by a clinical genetics laboratory in order to identify variants eligible for return. Participants are re-contacted without disclosing the specific research result and a second sample is requested that can be confirmed with clinical testing. Upon confirmation, the result is communicated, most often by a genetic counselor or physician associated with the study, who then assists the participant in pursuing appropriate referrals. The complexity and costs of implementing this gRoR template are intimidating to most researchers, and detailed logistical data, time utilization and costs from sites conducting gRoR have not been previously reported. In this report, we provide a comprehensive overview of one gRoR protocol within the biobank of a large healthcare system and present detailed data on consent processes, initial research laboratory analysis and verification, recontact efforts, clinical laboratory confirmation of research findings, results disclosure and clinical referral among biobank participants, as well as the effort and costs required to carry out such a protocol.

## MATERIALS AND METHODS

### Protocol Design

The Mass General Brigham Biobank (MBG Biobank) is a research biorepository in an academic medical center linked to electronic health records (EHR).^26^ The protocol was approved by the Mass General Brigham Institutional Review Board (IRB). An MGB Biobank Return of Results Committee designed the protocol for recontact and disclosure of genomic results with input from participant stakeholders and the IRB. The consent and disclosure process followed an incremental disclosure protocol in which participants were consented upon biobank enrollment with the explicit understanding that their DNA would be analyzed for research, and that they might be recontacted if “medically important” results were discovered (S1). The option to decline recontact was not available if participants consented to enroll in the biobank.

The genes selected for gRoR were the 59 genes in the 2nd version of the American College of Medical Genetics and Genomics (ACMGv2) recommended list to be evaluated for return of secondary findings during indication-based sequencing.^27; 28^ In these genes, only pathogenic or likely pathogenic (PLPV) variants classified according to the ACMG/AMP criteria met our reporting criteria for return (Figure 1).^29^

**Figure 1.**
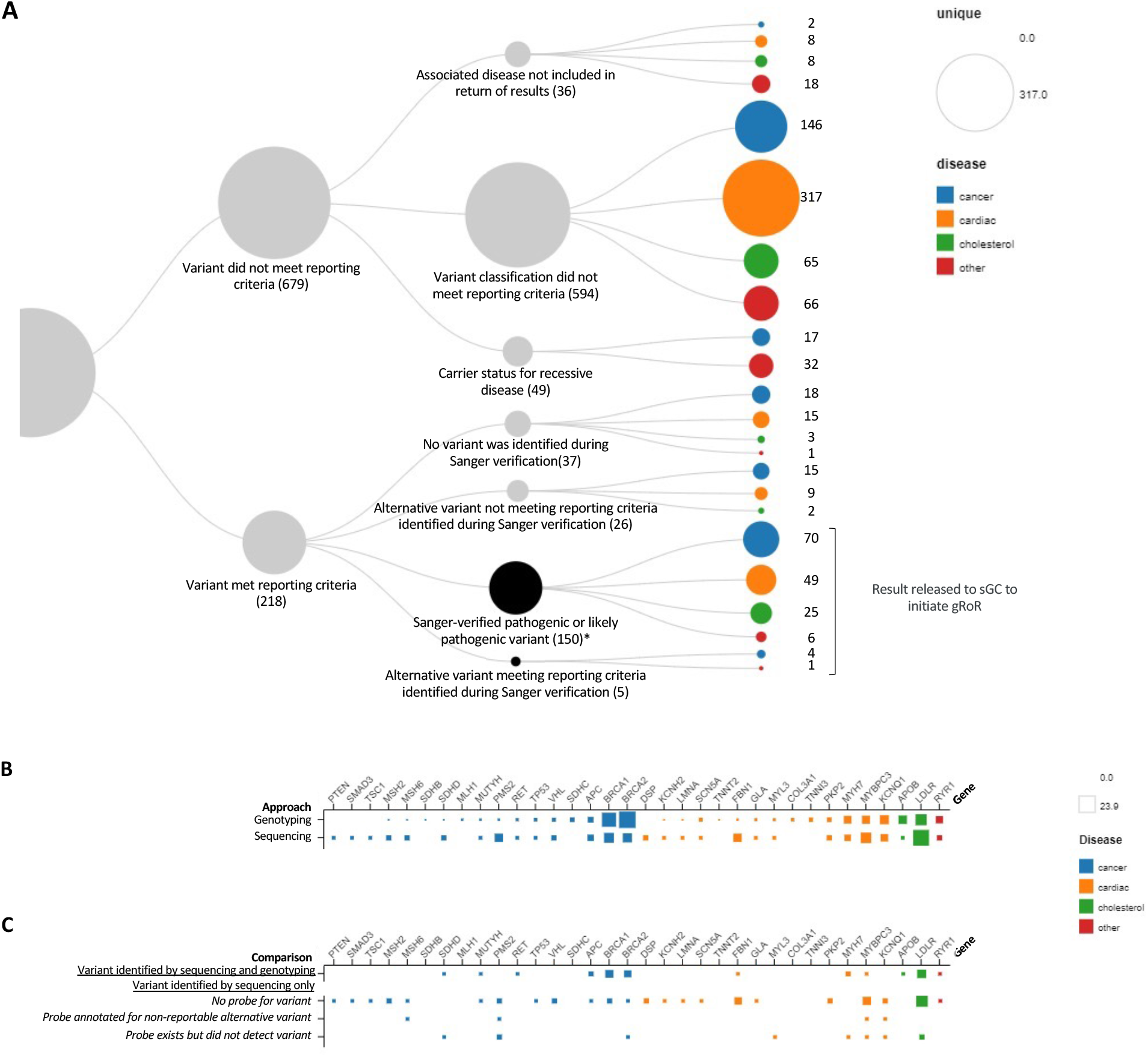
Interpretation of research data and yield per platform, disease area and gene. (A) Flowchart of research interpretation of unique variants revealed in genotype (GT) data. Among 36,417 participants whose DNA was analyzed by GT, 218 unique variants initially met criteria for return, out of which 155 were replicated or revealed to have alternative reportable variants through Sanger verification of a second (non-CLIA) research sample. *This includes 3 variants that were downgraded after initiation of the gRoR process. Colors correspond to disease areas: cancer (blue), cardiac (orange), cholesterol metabolism (green) and other actionable conditions (red). (B) Among the 36,417 participants whose DNA was analyzed by GT and the 3,263 participants whose DNA was analyzed by genome sequencing (GS), the percentage of cases per gene is represented by the size of the squares showing the differences in relative frequency of genes by each platform, using the same color coding as above. (C) Among the 3,263 participants who were additionally analyzed by GS, squares represent the percentage of variants in each gene that were either also identified by GT or identified by sequencing only, along with the reasons that the variant was missed by GT for each gene.

For those participants in whom a PLPV was discovered in an ACMGv2 gene, a disclosure team of 1 part-time study-supported genetic counselor (sGC) and 2 part-time study-supported medical geneticists (sMG) organized and implemented the workflow, notified participants, collected samples for CLIA confirmation and facilitated final disclosure and clinical follow-up (Figure 2). Participants who had verified PLPVs in one of the genes on the ACMGv2 gene list, who were still living and who did not have prior personal knowledge or EHR record of the variant were considered eligible.

**Figure 2.**
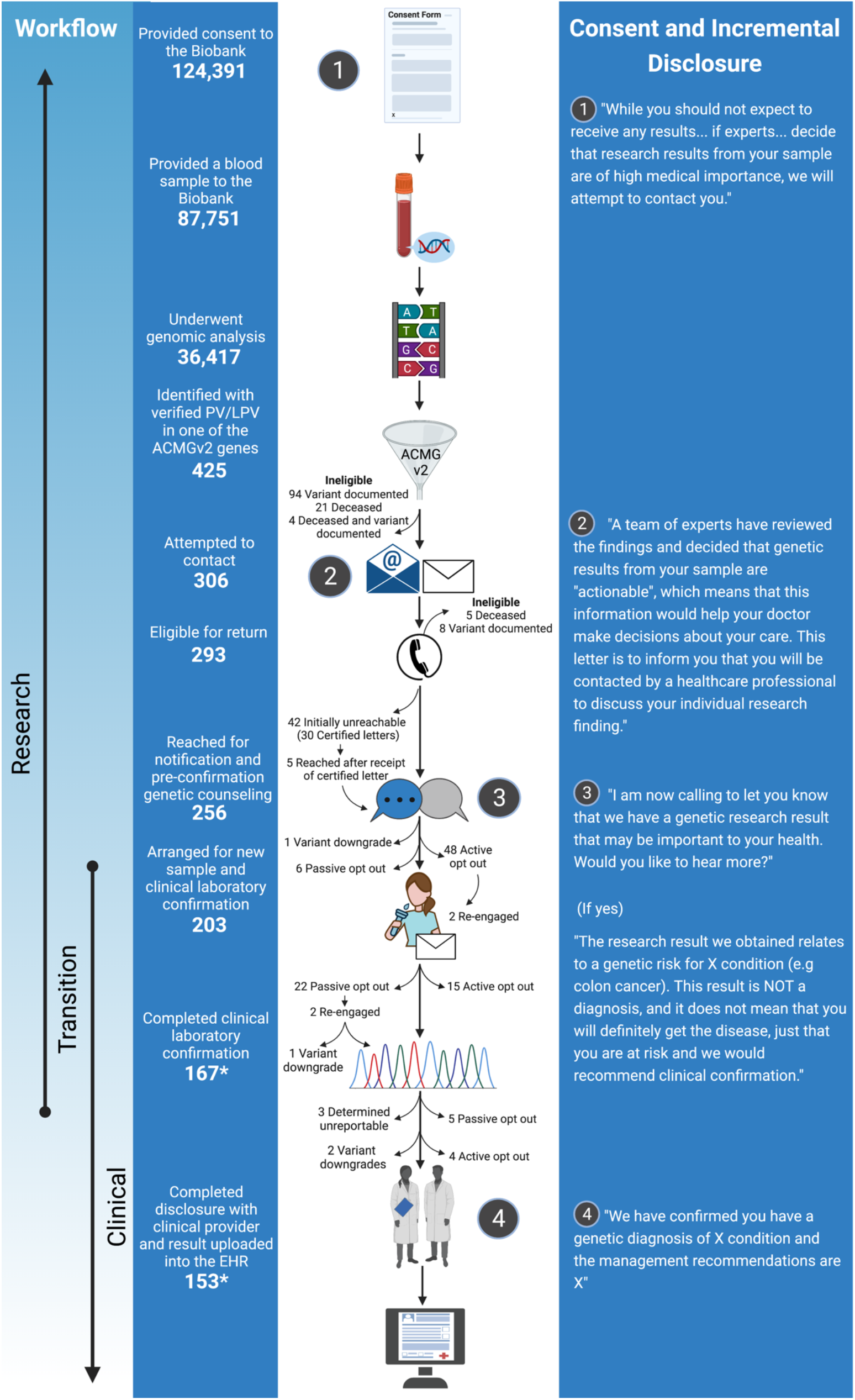
Participant flow through the biobank incremental disclosure gRoR process. *This number includes 10 participants that have elected to proceed with gRoR and are in progress but have not completed it.

Eligible participants were sent a letter alerting them to an actionable DNA finding without specifics, followed by a sGC call, with letters and calls repeated for up to 7 total contact attempts (S2, S3). If the participant was never reached but had a known address, a final certified letter was sent. Additional phone calls were made as needed and in response to participant requests and returning missed calls, and contact attempts were logged in a REDcap database (Figure 3, S4). In January, 2001 the number of letters sent was reduced from 4 to 2 letters, a first letter and a final certified letter. After reaching a participant, the sGC followed a phone script (S3) reminding them of their participation in the biobank, reiterating that a DNA result of medical importance had been identified and asking if they wished to hear more. If they agreed to hear more, the sGC described the specific condition associated with the genetic finding (e.g. colon cancer), but did not specify the gene or variant (S3), and counseled the participant about the implications of gRoR while collecting a brief medical and family history. Participants were given the opportunity to continue, or opt out, of a Clinical Laboratory Improvement Amendments (CLIA) approved laboratory confirmation (CLC) and results disclosure (Figures 2 and 3, S4). For participants who wished to continue, a clinical saliva or blood sample was collected and accessioned by the CLIA and CAP-approved MGB Laboratory for Molecular Medicine (LMM), and variants were confirmed by Sanger sequencing in a CLIA compliant workflow. Laboratory results were finalized into a clinical report (S5) and shared with the sGC who assisted in identifying a provider (a medical geneticist, disease specialist, or their own PCP if requested) to handle disclosure in a conventional clinical appointment to ensure appropriate medical follow-up. MGB specialists or the sMGs returned results if the participant’s provider was unwilling to do so. The cost of CLC genetic testing was covered by the study, but the disclosure visit was considered a clinical service to be covered by a participant’s own medical insurance, and was scheduled with a physician who was prepared to contextualize the CLC finding, document the result in the official medical record and make further referrals and follow-up as medically indicated (Figure 2). The responsibility of the research team was considered to have ended when the clinician disclosed the clinical report to the participant.

**Figure 3.**
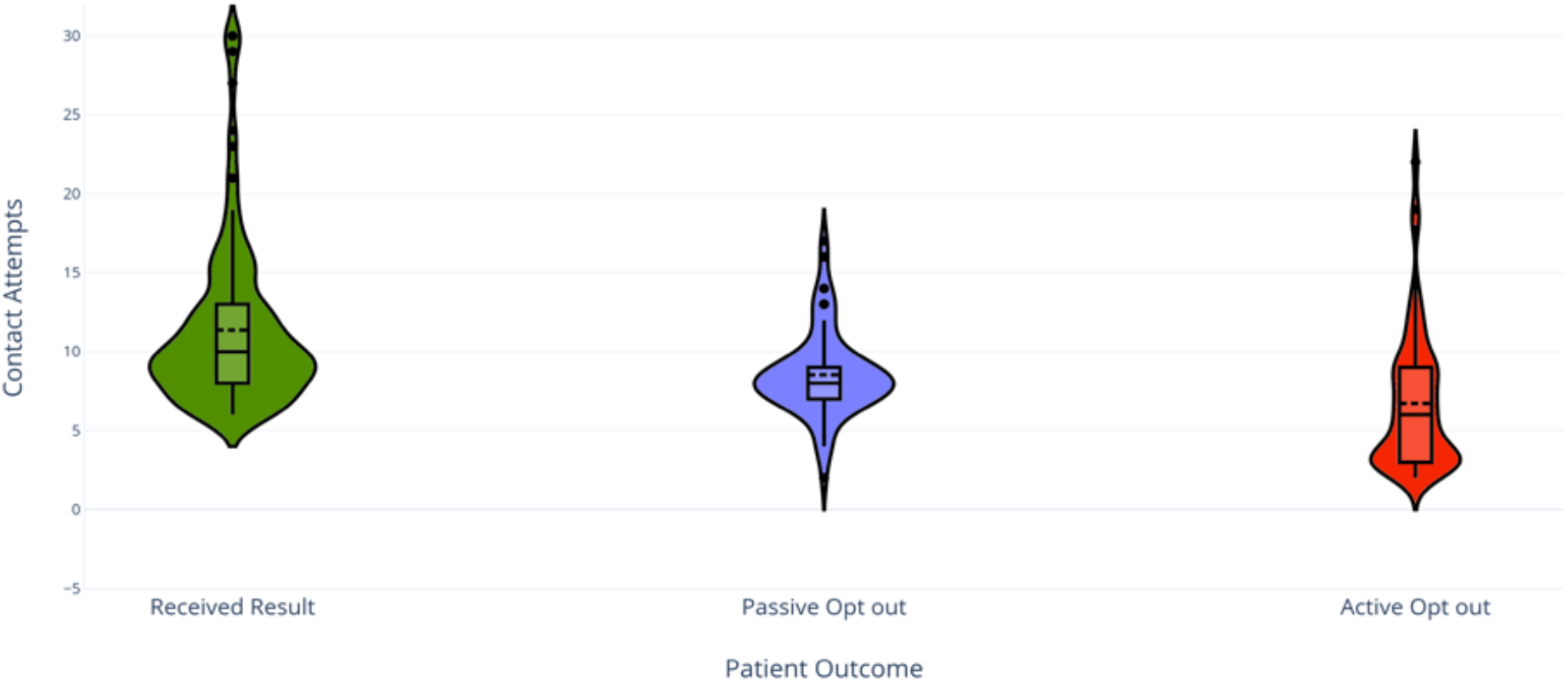
Number of contacts and contact attempts needed for each participant outcome. Participants are grouped into three kernel density plots that show the range of contact attempts needed to successfully disclose a result to participants (green) or to reach active (red) or passive (purple) opt out of the gRoR process. Also shown within each shape are box plots and interquartile ranges, with mid-plot solid line indicating the mean, and mid-plot dashed line indicating the median. Outliers in each violin plot are indicated by dots and represent situations in which multiple contacts (“please call me back”, “I’d like to think about it further”) were needed before the participant agreed to progress to results disclosure, ceased responding (passive opt out) or finally declined to proceed (active opt out). This figure excludes in-progress participants.

### Laboratory Methods

#### Genotyping arrays

Genotyping of 36,417 MGB Biobank samples utilized one of three versions of the Illumina (San Diego, CA) Infinium Multi-Ethnic Genotyping arrays that were developed to capture both variation among a diversity of genetic backgrounds and functional variation within the genome. The three versions included: 1) a pre-release version developed by the Multi-Ethnic Genotyping Array Consortium (MEGA), 2) an expanded version of the pre-commercial Expanded Multi-Ethnic Genotyping Array (MEGAEX), and 3) the final commercial version of the Multi-Ethnic Global (MEG). A substantial number of probes from MEGA and MEG were re-designed in later versions due to quality and probe re-synthesis issues. Therefore, for both pre-commercial arrays the analyzed probes were limited to the content of the final MEG array. Additionally, for each array the probe coordinates were remapped based on the TopGenomicSeq in the manifest provided by Illumina to more accurately determine probe location for downstream annotations.

#### Sample Processing

The initial 4924 samples assayed using the MEGA array were genotyped at Illumina, while genotyping using the MEGAEX and MEG arrays were conducted internally at the MGB Biobank Genomics Core. For all samples, quantification to assess the concentration of double-stranded DNA was conducted using picogreen, and 200-400 ng of genomic DNA was amplified using a whole genome amplification process. Genotyping was then performed following the same procedures using the Illumina-recommended protocol. Quality control (QC) of the genotyping arrays was carried out by looking at the Controls Dashboard within Genome Studio. These controls monitor internal spike in probes at various points of the process and allow the QC of sample dependent and sample independent processes.

#### Genotyping processing

For analysis, batch sizes targeting 5,000 subjects were created to enable quicker return of actionable results rather than waiting for all genotypes to be generated. For each batch of data, the GTC files generated in Illumina LIMS were converted to PED file format using a customized script based on Illumina provided code that tracks array annotations and specimen annotations. During this conversion from GTC files to PED, pre-defined probes that underperformed (<95% call rate) or were not able to be mapped to GRCh37 were removed.^30^ All samples required an overall call rate >99% as calculated by Illumina LIMS using a cluster file specific for each array version. Data for subjects in which a gender mismatch was identified and not resolved or data for a participant that had subsequently withdrawn from the study were removed from analysis. The datasets for the remaining subjects were converted to individual vcf files.

#### Genomic sequencing

A subset of 2349 genotyped individuals were sequenced for a limited set of genes as part of the Electronic Medical Records and Genomics (eMERGE) III program.^31^ Further, a set of 914 additional individuals who self-reported as Hispanic or Latino, Black or African American or other in the MGB EHR were prioritized for analysis from exome sequencing data. Briefly, exome sequencing was run at the Broad Institute of Harvard and MIT using a custom capture library from TWIST Biosciences (approximately 37Mb target) with sequencing on the Illumina NovaSeq using 150 bp paired reads. The hybrid selection libraries met or exceeded 85% completeness of exonic targets at 20x, which is comparable to approximately 55x mean coverage. Exome sequences were aligned to GRCh38, joint-called using the Genome Analysis ToolKit (GATK) across the biobank cohort, and converted back to GRCh37 coordinates for interpretation.

#### Variant interpretation

Variants were filtered to a list of 59 genes included in ACMGv2 (S6).^27; 28^ For comparisons in the paper, we divided the ACMGv2 genes into the following categories: cancer (*SDHD, SDHAF2, SDHC, SDHB, STK11, PTEN, MEN1, MUTYH, MLH1, MSH2, MSH6, PMS2, APC, BRCA1, BRCA2, RET, BMPR1A, SMAD4, TP53, RB1, VHL, WT1*); cardiac disease (*MYH7, TPM1, PRKAG2, TNNI3, MYL3, MYL2, ACTC1, TMEM43, DSP, PKP2, DSG2, DSC2, SCN5A, RYR2, LMNA, MYBPC3, GLA, TNNT2, KCNQ1, KCNH2, COL3A1, MYH11, ACTA2, TGFBR1, TGFBR2, SMAD3, FBN1*); familial hypercholesterolemia (*APOB, LDLR, PCSK9*); and other actionable diseases (*ATP7B, RYR1, CACNA1S, OTC, TSC1, NF2, TSC2*). The variant calls within the set of 59 genes were annotated using multiple data sources, including Alamut (Alamut Batch, SOPHiA GENETICS, Lausanne, Switzerland), the Genome Aggregation Database (gnomAD),^32^ ClinVar,^33^ the Human Genome Mutation Database (HGMD),^34^ and the GeneInsight Suite (Sunquest, Tucson AZ).^35^ The annotated variants were filtered using the GeneInsight Suite to find: 1) variants previously identified as disease causing by the MGB LMM, 2) variants classified as P/LP within ClinVar with a minor allele frequency (MAF) <5.0%, 3) variants classified as a disease-causing mutation (DM) in HGMD with a MAF <5.0%, and 4) loss-of-function variants (nonsense, frameshift, canonical splice-site, and initiating methionine variants) with a MAF <1.0%. Variants of Uncertain Significance (VUS) were not reported, however some variants were downgraded to VUS over the course of the study (Figure 2, Table 2, S6). Clinical variant classification was carried out in accordance with the criteria set by the guidelines by the ACMG and the Association of Molecular Pathology,^29^ with disease specific modifications as recommended by the Clinical Genome Resource Expert Panels.^36^ Confirmation of PLPVs was conducted on the research sample prior to initiation of gROR to ensure variant calling accuracy; for genotyping results, research confirmations were not conducted once the variant call was determined to be high confidence or a clear false positive. Only PLPVs associated with disorders listed in the ACMGv2 gene list ^28^ were returned to participant, and if seen with the following genotypes: heterozygous, homozygous, or biallelic PLPVs for autosomal dominant conditions; homozygous or biallelic PLPVs for autosomal recessive conditions; and heterozygous, homozygous, hemizygous, or biallelic PLPVs for X-linked conditions (Figure 1).

### Electronic Health Record Review

We reviewed the EHR for medical and family history and assessed whether, prior to disclosure, participants met published criteria from the National Comprehensive Cancer Network (NCCN) for genetic testing for colorectal and hereditary breast and ovarian cancer predisposition,^37-40^ or professional society/expert guidelines for other genes leading to cancer predisposition, where NCCN guidelines were not available;^40; 41^ as well as modified Dutch Lipid Clinic Network (DLCN) criteria for familial hypercholesterolemia (FH) (not awarding points for discovery of the genetic variant).^42; 43^ We then assessed whether obtaining additional targeted personal and family history at the time of notification and disclosure would have changed that participant’s eligibility for recommended clinical genetic testing (Figure 4).

**Figure 4.**
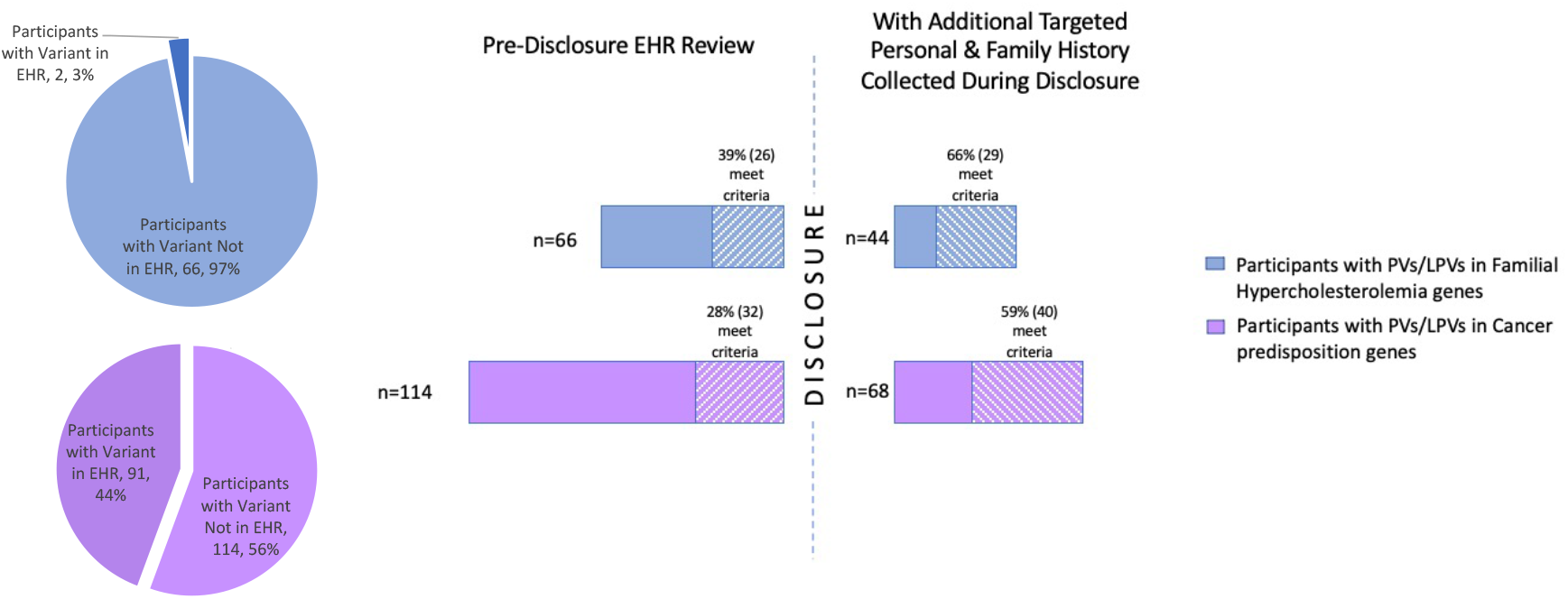
Electronic health record (EHR) review of those meeting professional guideline criteria for clinical genetic testing. EHR were reviewed for participants with PLPVs in 3 familial hypercholesterolemia (FH, blue) genes and 22 cancer predisposition genes (purple). Pie charts reveal the percentage of individuals whose PLPV was previously documented in the medical record. Chart reviews were performed using NCCN guidelines or other established expert criteria for cancer predisposition syndromes and the Dutch Lipid Clinic Networks guidelines for FH. The bar graphs show the percentage of participants whose PLPV variant was not previously documented in the EHR, but who nonetheless met expert criteria for ordering genetic testing on the basis of EHR review alone (pre-disclosure EHR review) and the percentage of participants who met expert criteria for ordering genetic testing on the basis of EHR review and additional personal and family history gathered from the participant in the process of disclosure.

### Surveys

Participants who opted in for gRoR were sent surveys by email, or if requested by mail, at Baseline (after notification but before clinical disclosure), 1 month post genomic results disclosure and 6 months post disclosure to assess their decisional regret with gRoR using a published 5-item decision regret scale.^44^ Responses at 1 and 6 months post disclosure were converted to a 0-100 score based on scale instructions, with higher scores indicating greater regret about that decision. Scores above 50 were considered to indicate overall regret (i.e., a tendency to agree with statements such as, “I regret the choice that was made”).

### Interviews

Among the 65 active and 31 passive decliners, a convenience sample of 51 (34 active and 17 passive) decliners were contacted to ascertain reasons for declining (S7). Twenty-four (17 active and 7 passive) decliners verbally consented to semi-structured phone interviews. Interviews, lasting 5-25 minutes, were audio recorded, transcribed, and uploaded into NVivo 12 (QSR International, Melbourne, Australia). A codebook developed by two coders (MU, JS) was used to perform consensus coding on transcripts using thematic analysis. Codes were grouped according to similar themes, representing reasons for declining.

### Budget Impact and Cost Analysis

A time and budget impact analysis was conducted to estimate the incremental research costs to incorporate gRoR using this protocol, including laboratory verification of previously genotyped and sequenced samples, recollection and CLC of new samples, as well as estimated salaries for program oversight and staffing (Figure 5).^45^ Laboratory personnel costs and effort were estimated for generating genetic research results and for CLIA confirmation, while material costs were actual. Efforts by the team to review medical records, inform individuals about the research completed finding, coordinate confirmatory testing and clinical disclosure sessions were estimated using a modified micro-costing approach^46^ where time estimates of all logged contacts were multiplied by median national hourly costs for the relevant personnel and adjusted for wage inflation.^47^ Fixed costs included office space and personnel costs, including monthly meetings of the 19-member MGB Biobank Return of Results Committee during a 46-month period, including monthly effort for Committee leadership, and 3 months of Committee time to establish the gRoR pipeline (e.g., protocol creation and IRB review). Cost analyses are presented in 2021 U.S. dollars and included the costs associated with obtaining a second DNA sample and performing CLC of the second sample. Costs of the research GT/GS, the medical appointments for confirmatory variant disclosure, and subsequent costs for patient management were not included in these estimates.

**Figure 5.**
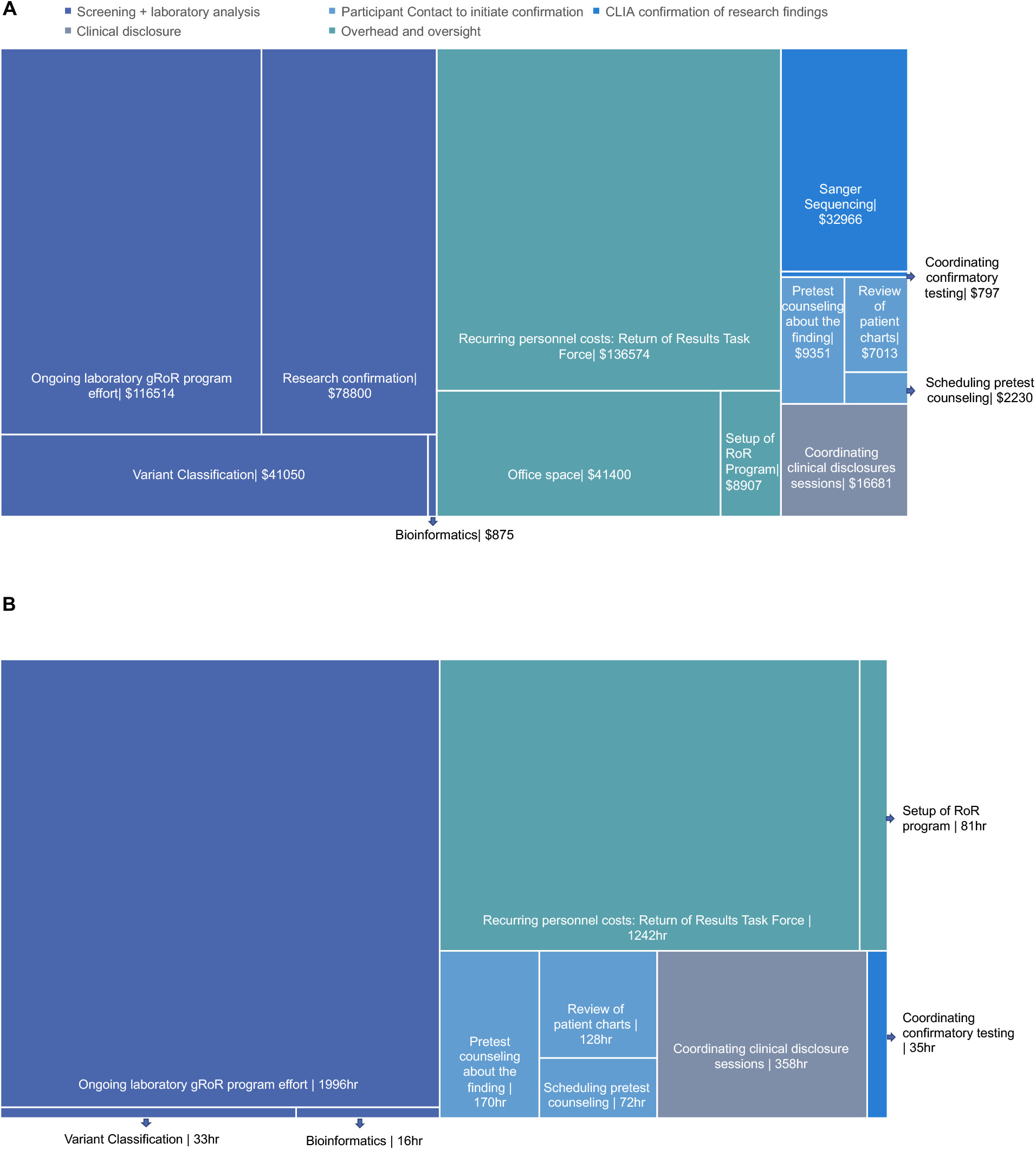
Cost and Time Impact Analysis of gROR to MGB Biobank participants. Panel A shows a treemap of research cost (in 2021 US dollars) whereas panel B shows a treemap of research personnel time (in personnel hours) invested in analysis and subsequent gRoR across all Biobank participants. Research based confirmation and CLIA-based Sanger sequencing confirmation are accounted for as reagent costs only, hence do not have a time associated with them, whereas office space is accounted for as a fixed cost that did not change for the duration of the gRoR process, hence these metrics are not indicated in Panel B.

## RESULTS

### Participants

Between July 1, 2010 and March 31, 2021, the MGB Biobank enrolled 124,391 individuals, of whom 87,751 provided a blood sample. Beginning in 2015, DNA samples on 36,417 participants were genotyped (GT) using one of Illumina’s Multi Ethnic Global arrays (Table 1, Figure 2). Two subsets of the samples that underwent GT also underwent genomic sequencing (GS): these samples were from (1) a cohort of 2,349 participants in whom a limited set of medically actionable genes was sequenced as part of the Electronic Medical Records and Genomics (eMERGE) III program^31^ and (2) a cohort of 914 additional underrepresented minorities (Black or African American, Hispanic or Latino, or Other) that were prioritized for analysis of exome sequencing. Table 1 shows the demographics of the participants in the MGB Biobank, those whose DNA was analyzed, those in whom returnable findings were identified, those who were eligible for results return and those in whom results were disclosed or in whom disclosure is underway. Because genetic analysis and interpretation lagged behind consent and enrollment, participants were consented an averaged 3.4 years (range 1.8-8.9 years) before they were contacted for gRoR.

**Table 1.** Demographics of Biobank Participants*. *Plus-minus values are means ± SD. Percentages may not total 100 because of rounding.

| Characteristic | Biobank participants<br>N=124,391 | DNA was analyzed<br>N=36,417 | Returnable finding identified<br>N=425 | Eligible for return<br>N=293 | Result disclosed or disclosure in progress<br>N=153 |
| --- | --- | --- | --- | --- | --- |
| Female sex – no. (%) | 70,612 (56.8%) | 19,713 (54.1%) | 232 (54.6%) | 149 (50.9%) | 79 (51.6%) |
| Age - yr | 56.1 (±17.7) | 59.9 (±17.1) | 59.3 (±16.2) | 59.0 (±16.7) | 58.2 (±15.6) |
| **Race/Ethnicity – no. (%)** |  |  |  |  |  |
| Non-Hispanic White | 103,587 (83.3%) | 30,302 (83.2%) | 361 (84.9%) | 245 (83.6%) | 134 (87.6%) |
| Non-Hispanic Black | 5,652 (4.5%) | 1,758 (4.8%) | 19 (4.5%) | 14 (4.8%) | 4 (2.6%) |
| Non-Hispanic Asian | 3,662 (2.9%) | 815 (2.2%) | 7 (1.6%) | 6 (2.0%) | 3 (2.0%) |
| Hispanic | 6,394 (5.1%) | 2,227 (6.1%) | 27 (6.4%) | 19 (6.5%) | 7 (4.6%) |
| Unknown/Other | 5,095 (4.1%) | 1,315 (3.6%) | 12 (2.8%) | 9 (3.1%) | 5 (3.3%) |

### Participant Contact

We tabulated the number of contacts required to notify participants of the research results, as well as the number of participant and provider contacts required to arrange for CLC and disclosure among those who elected disclosure, those who eventually opted out at any point (active opt out) and those who were reached but ceased responding to our calls (passive opt out) (Figures 2 and 3, and Table 2). Of the 425 participants identified with actionable variants, we found 293 who were eligible for return after EHR review and initial contact attempts. We reached 256 (87%) of these for result notification and pre-confirmation genetic counseling, confirmatory sample collection was initiated for 203 (69%) individuals (192 saliva kits and 11 blood draws), results were confirmed by CLC and disclosed to 143 (49%) participants, and 10 are currently in the process of confirmation (Figure 2, Table 2).

**Table 2:** Breakdown of participants with a lab reportable variant discovered by result type.

|  | % of participants out of total with a lab reportable finding |  |  |  |  |
| --- | --- | --- | --- | --- | --- |
|  | Total =425 | Cancer =209 | FH= =75 | Cardiac= 122 | Other=19 |
| **Variant previously documented** | 24.9%<br>(n=106) | 43.1% (n=90) | 2.7% (n=2) | 9.8% (n=12) | 10.5% (n=2) |
| **Deceased** | 7.1% (n=30) | 8.1% (n=17) | 8% (n=6) | 4.1% (n=5) | 10.5% (n=2) |
| **Eligible for return** | 68.9%<br>(n=293) | 50.7% (106) | 89.3% (n=67) | 86.1%<br>(n=105) | 78.9% (n=15) |
| **Result disclosed** | 33.6%<br>(n=143) | 29.7% (n=62) | 26.7% (n=20) | 41% (n=50) | 57.9% (n=11) |
| **Variant downgraded during gROR** | 0.9% (n=4) | 0% (n=0) | 1.3% (n=1) | 2.5% (n=3) | 0% (n=0) |
|  | % of participants out of total eligible for gROR |  |  |  |  |
|  | Total =293 | Cancer =106 | FH=67 | Cardiac= 105 | Other= 15 |
| **Unreachable** | 12.6% (n=37) | 10.4% (n=11) | 17.9% (n=12) | 12.4% (n=13) | 6.7% (n=1) |
| **Reached** | 87.4%<br>(n=256) | 89.6% (n=95) | 82.1% (n=55) | 87.6% (n=92) | 93.3% (n=14) |
|  | % of participants out of total reached |  |  |  |  |
|  | Total =256 | Cancer =95 | FH=55 | Cardiac= 92 | Other= 14 |
| **Opted out of return** | 37.5% (n=96) | 31.6% (n=30) | 52.7% (n=29) | 37.0% (n=34) | 21.4% (n=3) |
| **Active opt out** | 25.4% (n=65) | 27.4% (n=26) | 34.5% (n=19) | 19.6% (n=18) | 14.3% (n=2) |
| **Passive opt out** | 12.1% (n=31) | 4.2% (n=4) | 18.2% (n=10) | 17.4% (n=16) | 7.1% (n=1) |
|  | % of participants out of total in which sanger confirmation attempted |  |  |  |  |
|  | Total n=162 | Cancer= 66 | FH=28 | Cardiac= 57 | Other= 11 |
| **Sanger confirmed variants** | 98.1%<br>(n=159) | 98.5% (n=65) | 92.9% (n=26) | 100% (n=57) | 100% (n=11) |
| **Variant not reportable after clinical Sanger sequencing** | 1.9% (n=3) | 1.5% (n=1) | 7.1% (n=2) | 0% (n=0) | 0% (n=0) |

### Research Laboratory Findings and Verification

Variants from both GT and GS were filtered and classified to identify PLPVs in the ACMGv2 genes for possible gRoR. Initial inspection of GT samples indicated a high proportion of false positive calls, so a Sanger verification step was performed on samples that yielded PLPVs by GT prior to participant contact. This verification step determined that 28.9% (63/218) of unique variants and 44.9% (302/673) of the samples were analytic false positives (Figure 1A). As expected, GS showed very high rates of verification.^48^ A total of 425 unique participants had a PLPV identified in Sanger-verified GT or GS (Figure 2). PLPVs among the ACMGv2 genes were found in 1.0% (368/36417) of participant samples that underwent Sanger-verified GT and 2.5% (82/3263) of those that also underwent GS. Detection of PLPVs in the GT data was limited to those variants/conditions present on the array, as compared to the unbiased GS data (Figure 1B). Among those participants whose samples underwent both GT and GS, there were 79 unique variants in 82 participants identified by GS, but 58 of these variants in 59 participants (72.0%) were missed by GT due to the absence of a probe on the array (45 unique variants) or due to poor performing or incorrectly annotated probes (13 unique variants) (Figure 1C).

### Transitioning Participants into the Clinical Workflow

Of 425 participants initially identified with PLPVs in the ACMGv2 genes, EHR review or phone notification revealed that 30 (7.1%) were deceased, 106 (24.9%) were previously known to have the variant, including 4 that fell in both categories. A total of 256 eligible participants were reached for pre-confirmation counseling by the sGC, including 4 individuals whose variants were downgraded during the gRoR process and 3 individuals whose variants were determined to be unreportable during clinical confirmation (Figure 2, Table 2). Between 2 and 12 contact attempts were required to reach each participant for result notification and counseling, and between 4 and 28 additional contact attempts with participants and providers were needed to facilitate final results disclosure (Figure 2, 3, S4). Of the 256 participants who were alerted that they carried medically important DNA change, there were a total of 65 active and 31 passive decliners. Four initial decliners re-engaged in the disclosure process, for a total of 96 participants who declined and an overall decline rate of 37.5% (Figure 2, Table 2). Comparing those who declined by category of underlying condition, there were 30 of 95 participants reached (31.6%) who declined after being alerted that they carried a variant for increased cancer risk, 29 of 55 participants reached (52.7%) who declined after being alerted that they had a variant for a hereditary high cholesterol disorder, 34 of 92 participants reached (37.0%) who declined after being alerted that they had a variant for a (non-FH) heart condition and 3 of 14 participants reached (21.4%) who declined after being alerted that of a variant that would cause an abnormal reaction to surgical anesthesia (referring to *RYR1)* (Table 2). A subset of the decliners consisting of 34 active and 17 passive decliners, were contacted to ascertain reasons for declining, with 17 and 7, respectively, completing a qualitative interview (S7). The most common reasons for declining confirmatory testing were that individuals perceived their genetic results to be irrelevant (largely because they were already aware that they had the associated phenotype) or that they had more pressing medical concerns (S7). None of the participants who received notice of a medically important finding expressed distress about being alerted for potential gRoR or about the subsequent process of disclosure. Among those who elected to proceed with clinical confirmation and disclosure, it took an average of 88 days (median 56 days) from completed sGC notification to clinical result disclosure. Factors impacting this were how quickly participants provided a clinical sample for confirmation, time to generate the laboratory report and disclosure appointment scheduling.

### Comparison to Established Clinical Criteria for Genetic Testing

The EHR was reviewed for 418 participants (the total with a variant identified excluding those downgraded during gRoR (n=4) and those not reportable after Sanger confirmation (n=3)). Of those living and deceased participants who were found to have PLPVs in the ACMGv2 genes, 319/418 (76.3%) did not have the variant previously documented in their EHR. We reviewed the EHR for documented medical and family history and assessed whether, prior to disclosure, 180 participants without documentation of prior genetic testing met available expert criteria to prompt genetic testing for their condition from the National Comprehensive Cancer Network (NCCN) for genetic testing for cancer ^37; 38; 40; 41^ or the Dutch Lipid Clinic Network (DLCN) criteria for FH, without awarding points for research discovery of the PLPV.^42; 43^ Among participants without documentation of prior genetic testing, 32/114 (28%) with PLPVs in cancer predisposition genes fulfilled NCCN guidelines for genetic testing and 26/66 (39%) of those with PLPVs in FH genes were considered “likely” to have FH by DLCN criteria based upon EHR review alone (Figure 4). After obtaining additional family history at the time of notification and disclosure in 112 of the 180 participants, these proportions increased to 40/68 (58.8%) for NCCN criteria and 29/44 (65.9%) for DLCN criteria (Figure 4.).

### Assessment of Decisional Regret

A decision regret scale^44^ was administered as part of a larger survey at 1 and 6 months. Participants who completed the entire protocol and had their research result clinically confirmed and disclosed were asked how they felt about their decision to enroll in the study and receive results. At 1 month following disclosure, 57/111 (51.4%) responded to the survey, and only one individual scored in the range that suggested regret. The mean score was 8.8 on the 0-100 scale (in which higher scores indicate greater levels of regret), lower than observed in other studies of genetic disclosure to biobank populations^49^. At 6 months, 50/95 individuals (52.6%) responded to the survey with a mean score of 10.8 on the same scale, with 1 individual (a different individual than the 1-month respondent, who did not complete a 6-month survey) scoring in the range that suggested regret.

### Time and Budget Impact Analysis

Total costs for gRoR efforts using our protocol were estimated at $493,258, including $237,239 (48.1%) for screening and laboratory analysis, including initial verification and eventual CLC, and $136,574 (25.0%) for program oversight (Figure 5). Spread across the entire cohort of persons whose DNA was analyzed and the duration of the gRoR effort in the biobank, this represented approximately $14 per participant and approximately $129,000 per year. Genetic counselors and research assistants devoted 370 hours from May 2017 through March 2021 contacting participants about their result, 35 hours coordinating confirmatory testing and 358 hours coordinating clinical appointments for disclosure and subsequent care. Amortized across the 153 clinical disclosure sessions, each participant who eventually received disclosure in the clinical domain required 5.0 hours of time by the sGC and research assistants and cost the overall research team and associated laboratory approximately $3,224.

## DISCUSSION

In this report, we describe the consent, recontact, analysis, yield, effort and cost involved in analyzing research results for actionable genomic findings, confirming and disclosing these findings and transitioning participants who learn these findings into clinical care. Our gRoR protocol is not proposed as a criterion standard for how gRoR should take place, but it provides details and insights that may assist other investigators in designing their own gRoR protocols. In particular we document that 76.3% of individuals who carried actionable variants were unaware of this, and that between 59-66% of those met available professional guidelines to prompt genetic testing but had never been tested. While the vast majority of research participants across multiple studies claim they wish to be alerted to genetic findings of medical importance,^1-3^ 37.5% of those in our biobank who were contacted with such results actively or passively declined return of actionable results, despite numerous contact attempts. In addition, we document a cost of $14 per participant, above and beyond the initial research genotyping or sequencing, to cover our gRoR protocol, resulting in an average cost of $3,224 for each participant for whom gRoR was successfully completed.

Given limitations in participant understanding of consent,^50; 51^ it is extremely challenging to effectively educate and counsel every biobank participant about each of the rare conditions that might be revealed with gRoR. Our protocol utilized an incremental disclosure process for gRoR in which participants were not asked to finalize their willingness to receive genetic results upon enrollment, but rather were consented to recontact if the investigators discovered medically important findings. Various alternative models for gRoR consent (generic, staged, mandatory, tiered-layered-staged) have been proposed,^52-54^ but empirical data on these are scarce. Our approach shares some features with mandatory or staged consent models,^54; 55^ and has the advantage of reducing complexity during initial consent by moving the counseling and decision about additional information and disclosure to the time frame in which the participant would actually utilize the information, which in our biobank was up to 9 years after enrollment. The fact that more than one-third of our participants actively or passively opted-out of further disclosure once alerted to the fact that they carried an actionable genomic finding would suggest that the incremental disclosure process did not compromise participants’ freedom to decline full disclosure. And among those whom we could reach for follow-up inquiry, there was no distress recorded from those who opted out, nor any widespread regret among those who carried through to full disclosure.

Our data on the frequency of verified PLPVs among the ACMGv2 gene list in biobank participants are consistent with prior population screening efforts using this list that yielded a frequency of such variants of 1-1.5% among individuals who had been genotyped,^56^ and 2.6% among individuals who had been sequenced.^57-59^ Our data replicate and extend prior observations around the poor performance of GT as a potential tool for biobank gRoR or population screening.^56; 60-62^ Of the initial GT calls from over 36,000 participants from our biobank, nearly 45% of samples initially identified as carrying PLPVs were false positives. And in the subset of 3,263 participants who had both GT and GS, GT failed to detect a PLPV in 72.0% of the participants who were carrying GS-detected PLPVs. The comparison of GT and GS data also demonstrates a bias in identifying variants in certain genes and conditions that were not part of the array designs. Aside from common variants in *BRCA1* and *BRCA2*, variants indicating cancer predisposition were considerably less well-detected in GT as compared to GS. This bias may be different in other arrays such as the Global Screening Array (GSA) that was specifically designed for population-scale genomic studies around monogenic disease, but a study of over 5000 participants screened with a GSA array in Alabama revealed very similar figures for the overall yield and for the rate of analytic false positives.^56^ The limitations of GT are important to recognize as some healthcare systems and biobanks are already returning genomic results discovered through GT.^56; 63-65^

Returning genomic results from the MGB Biobank and other research studies reveals that expert guidelines to prompt genetic testing are not being followed in clinical care. Among all of our biobank participants identified to carry verified PLPVs, the molecular diagnosis was previously documented in the EHR for less than one-quarter of participants. This was particularly striking since over half of those participants with previously unrecognized PLPVs associated with heritable cancers or lipid disorders that have clear guidelines for treatment met published professional criteria for genetic testing.^37; 38; 40-43^ Expert clinical recommendations for genetic testing have not been translated into clinical care, as has been observed in other health systems.^23; 66-69^

It is well recognized that the anticipated logistical and financial burdens of gRoR may discourage research biobanks from considering gRoR.^70; 71^ Setting aside the cost of the original research genotyping or sequencing, and ignoring downstream medical costs that might be triggered by the disclosure of the finding, the design, oversight and implementation of our entire gRoR protocol, including laboratory verification of initial GT findings and coverage of CLC cost, was approximately $129,000/year over 4 years, representing about $14 per participant or $3,224 per participant in whom a verified and confirmed result was successfully disclosed. These figures contrast with $605 per participant-disclosure for gRoR for the return of 6 aortopathy genes^49^ and $750 per participant-disclosure for a subset of the ACMGv2 gene list in a pediatric biobank.^22^ The difference in cost estimates may be because those studies did not actively screen for variants unrelated to participants’ presenting diagnoses and omitted most overhead costs (34% of our total estimated costs). Our cost estimates did not include expenses to the healthcare system incurred during and after clinical disclosures, however, there is emerging evidence from economic models that genomic risk information may be cost-effective.^72-74^

Resampling participants for CLC is a routine part of gRoR in most US environments because research genotyping and sequencing is typically not conducted through a CLIA approved laboratory process that asserts quality control along the chain of custody and within the laboratory itself; and there have been widely accepted assertions by the Centers for Medicare and Medicaid Services (CMS) that laboratory results generated in a non-CLIA process should not be disclosed to individuals.^75^ But as shown in Figure 2 and 3 and Table 2 a substantial proportion of participants who were reached and informed that they carried a medically important variant actively or passively declined to complete the process, either before or after they submitted a second sample for confirmation of the research result. Some of these opt outs may have represented authentic decisions to avoid confronting a medical risk, but others may have represented insufficient motivation to overcome the barrier of multiple communication steps with study staff, or of submitting a new DNA sample.

There are a number of important limitations to this report. Our biobank recruited patients within an urban academic healthcare system. Like all gRoR models, ours depends upon the ability of biobank personnel to successfully recontact participants and biobanks that aggregate participant data from multiple sites would face a different set of challenges.^76^ Our interactions with participants through surveys and decliner interviews did not reveal regret over recontact and notification, but not all decliners were reached for interviews, not all who received a result completed a survey, and some that we did not reach could have been confused or distressed. As final disclosures were conducted in a clinical setting, this could present challenges to uninsured or underinsured patients. The proportions of Hispanic or Black participants, though consistent with the proportions of participants in the biobank, were small, so our findings may not be applicable to participants in racial or ethnic groups that have experienced disparities, and additional research is needed in these populations.

While it is sometimes difficult to achieve consensus on what constitutes actionable genomic findings, it is clear that this category is expanding,^77^ and that there will be increasing interest in, and demand for, gRoR. Although planning for gRoR in a research biobank can be complex, we hope the results of this study illuminate lessons learned that can be considered by other groups seeking to find the balance between conducting scientific research, preserving participant autonomy and privacy, and offering information that could reduce morbidity and mortality among those who have generously contributed their DNA for the benefit of science.

## Supporting information

COI disclosure forms

## Data Availability

Individual-level MGBB data are available from:
https://personalizedmedicine.partners.org/Biobank/Default.aspx, however, there are restrictions on this data which was accessed under IRB protocol 2009P002312 for this current study so are
not publicly available. Samples sequenced or genotyped as part of the eMERGE consortium are deposited in dbGAP https://www.ncbi.nlm.nih.gov/projects/gap/cgi-bin/study.cgi?study_id=phs001616.v1.p1 and https://www.ncbi.nlm.nih.gov/projects/gap/cgi-bin/study.cgi?study_id=phs001584.v2.p2.

https://personalizedmedicine.partners.org/Biobank/Default.aspx

https://www.ncbi.nlm.nih.gov/projects/gap/cgi-bin/study.cgi?study_id=phs001616.v1.p1

https://www.ncbi.nlm.nih.gov/projects/gap/cgi-bin/study.cgi?study_id=phs001584.v2.p2

## Supplemental data

S1: MGB Biobank consent form

S2: Letter informing participant of actionable result

S3: Phone script and communication guide

S4: Contacts needed for each participant outcome

S5: Example clinical report

S6: Sanger Confirmed P/LP Variants, Alternative Variants Identified During Sanger Confirmation, Variants Downgraded After Contact

S7: Decliner interview guide and Reasons for Decline

## Declaration of Interests

Dr. Weiss has received compensation from UpToDate

Dr. Smoller is a member of the Leon Levy Foundation Neuroscience Advisory Board and received an honorarium for an internal seminar at Biogen, Inc. He is PI of a collaborative study of the genetics of depression and bipolar disorder sponsored by 23andMe for which 23andMe provides analysis time as in-kind support but no payments.

Dr. Green has received compensation for advising the following companies: AIA, Genomic Life, Grail, Humanity, Kneed Media, Plumcare, OptumLabs, Verily, VibrentHealth; and is co-founder of Genome Medical.

## Acknowledgements

We thank the following who made this study possible: the biobank patient participants, the research and clinical teams, the Mass General Brigham Hospital System and the eMERGE network. Figure 2 was created using BioRender.com.

This work was primarily funded by NIH grant HG008685, with additional support from NIH grants HG008676, HG009173, HD090019, HG008676, HG009922, HL143295, OD026553, TR003201 and the Franca Sozzani Fund.

## Web Resources

Genotyping was then performed following the same procedures using the Illumina-recommended protocol (https://www.illumina.com/products/by-type/microarray-kits/infinium-multi-ethnic-global.html).

## Data and Code Availability

Individual-level MGBB data are available from:

https://personalizedmedicine.partners.org/Biobank/Default.aspx, however, there are restrictions on this data which was accessed under IRB protocol 2009P002312 for this current study so are not publicly available. Samples sequenced or genotyped as part of the eMERGE consortium are deposited in dbGAP https://www.ncbi.nlm.nih.gov/projects/gap/cgi-bin/study.cgi?study_id=phs001616.v1.p1 and https://www.ncbi.nlm.nih.gov/projects/gap/cgi-bin/study.cgi?study_id=phs001584.v2.p2.

## Notes

### Author Declarations

Mass General Brigham IRB protocols: 2009P002312 and 2015P000983

