## Supplementary Appendix for "Returning Actionable Genomic Results in a Research Biobank: Analytic Validity, Clinical Implementation and Resource Utilization"

S1: MGB Biobank Consent Form

S2: Letter Informing Participant of Actionable Result

S3: Phone Script and Communication Guide

S4: Contacts Needed for Each Participant Outcome

S5: Example Clinical Report

S6: Sanger Confirmed P/LP Variants, Alternative Variants Identified During Sanger Confirmation, Variants Downgraded After Contact

S7: Decliner Interview Guide and Reasons for Decline

**Subject Identification**

Protocol Title: **Partners HealthCare Biobank**
Principal Investigator: Scott T. Weiss, MD, MS

Description of Subject Population: Individuals seen at Partners HealthCare

#### 1. What is the purpose of this research?

Researchers at Partners HealthCare System (Brigham & Women's Hospital, Massachusetts General Hospital, and other Partners institutions) are studying how genes and other factors affect people's health and contribute to human disease. To perform this research, we are asking patients at Partners to participate in the Partners HealthCare Biobank (Partners Biobank or Biobank) with blood samples to be stored in a research tissue bank (the "Biobank"). Taking part in this research study is up to you. Your decision to participate will not affect your clinical care in any way. Your participation can help us better understand, treat, and even prevent diseases that affect your loved ones, your family's future generations, as well as the larger community.

If you have any questions before you sign this consent form or after you join the study, you can contact the Partners Biobank staff at 617-525-6700 from Monday - Friday 9a – 5p. The person in charge of the Partners Biobank is Scott T. Weiss, MD. If you want to speak with someone **not** directly involved in the study, contact the Partners Human Research Committee at 857-282-1900. There is also an attached fact sheet that expands on the consent form to provide definitions and additional information.

#### 2. What will happen in this study?

- You may be asked to donate a blood sample of up to 5 tubes (about 3 tablespoons). If blood samples for the Biobank are not collected today, they may be collected at a future time when you have a blood draw ordered by your doctor.We may also use blood, urine or tissue samples collected as part of your clinical care now or in the future that would otherwise be thrown away.
- We will also look at your medical records now and in the future to update your health information. We will store some of your health information in the study database.
- We may ask you to complete questionnaires about your health.
- We may contact you in the future to get additional information and ask if you are interested in joining other research studies.
- A notation that you are taking part in this research study may be made in your electronic medical record.

#### 3. For what type of research will my samples be used?

- We plan to do many types of biological and genetic research with your sample, for example, research on heart disease, cancer, diabetes, mental illness, or reproduction to name a few. Genetic research may include looking at some or all of your genes and DNA to see if there are links to different types of health conditions.

- We may create a "cell line" from your sample that will allow researchers to have an unlimited supply of your cells for research.
- We may use your cells to create pluripotent stem cells. This type of cell can be used to create different types of tissue, for example, heart, muscle, or lung cells. Your cells might be used in research that alters genes in the cells in order to study different diseases and normal healthy processes. Your cells might be mixed with other human cells, animal cells, or grown in lab animals like mice.
- We may share your samples and any cell lines that are created, your DNA sequence information, your health information, and results from research with other central tissue or data banks, such as those sponsored by the National Institutes of Health, so that researchers from around the world can use them to study many conditions.

#### 4. Will I get results of research done using my samples?

- You may receive a newsletter or other information that will tell you about the research discoveries from the Biobank. This newsletter will not identify you or describe any of your personal results. It may be sent via Patient Gateway email if you're on Patient Gateway, by unencrypted email to your email address if you gave us one, or by US mail to your home address.
- Generally, we will not return individual results from research using your samples and data to you or your doctor. Research using your sample is just a stepping stone in learning about health and disease. Most of the findings that come from studying your sample will not be relevant to your personal health. However, in the future, this may change.
- It is important to remember that research results are not always meaningful and are not the same as clinical tests. While you should not expect to receive any results from your participation in this research, if experts from the Biobank decide that research results from your sample are of high medical importance, we will attempt to contact you. In some situations, follow up testing might be needed in a certified clinical lab. You and your medical insurer may be responsible for the costs of these tests and any follow up care, including deductibles and co-payments.
- It is possible that you will never be contacted with individual research findings. This does not mean that you don't have or won't develop an important health problem.
- In the future, when research results are published, they may show that certain groups (for example, racial, ethnic, or men/women) have genes that are associated with increased risk of a disease. If this happens, you or others may learn that you are at increased risk of developing a disease or condition.

#### 5. What are the benefits to me? Will I be paid for my samples?

- You will not directly benefit from research conducted on your samples stored in the Biobank. We hope that research using the samples and information will help us understand, prevent, treat, or cure diseases.
- You will not receive payment for your samples. In some locations, your parking cost may be covered or you may receive a cafeteria voucher.

#### 6. What are the costs to me to take part in the research tissue bank?

There are no costs to you to participate in the Biobank.

#### 7. How are my samples and health information stored in the Biobank?

Staff at the Biobank will assign a code number to your samples and health information. Your name, medical record number, or other information that easily identifies you will not be stored with your samples or health information. The key to the code will be stored securely in a separate file.

#### 8. Which researchers can use my samples and what information about me can they have?

- Your coded samples and health information may be shared with researchers at Partners institutions. They may also be shared with researchers at non-Partners institutions or with for-profit companies that are working with Partners researchers. Your samples will not be sold for profit. We may use your samples and information to develop a new product or medical test to be sold. The hospital and researchers may benefit if this happens. There are no plans to pay you if your samples and information are used for this purpose.
- We will only share information that identifies you with researchers within Partners who have approval of the Partners ethics board. We will not share information that identifies you with researchers outside Partners.
- In order to allow researchers to share research results, agencies such as the National Institutes of Health (NIH) have developed secure banks that collect and store research samples and/or data from genetic studies. These central banks may store samples and results from research done using Partners Biobank samples and health information. The central banks may share these samples or information with other qualified and approved researchers to do more studies. Results or samples given to the central banks will not contain information that directly identifies you. There are many safeguards in place at these banks to protect your privacy.

#### 9. How long will the Biobank keep my samples and information?

We will store your samples and information indefinitely.

#### 10. Can I stop allowing my samples and information to be stored and used for research?

Yes. You can withdraw your permission at any time. If you do, your samples and your information will be destroyed. However, it will not be possible to destroy samples and information that have already been given to researchers. If you decide to withdraw please contact the Partners Biobank staff in writing. In case we need to contact you about medically important research results from your sample, please notify the Partners Biobank staff if your address changes.

| | |
|---|---|
| Partners Biobank<br>65 Landsdowne St, Room 142<br>Cambridge, MA 02139 | Phone: 617-525-6700<br>FAX: 617-768-8513<br> |

#### 11. What are the risks to me?

- The main risk of allowing us to use your samples and health information for research is a potential loss of privacy. We protect your privacy by coding your samples and health information.
- There is a risk that information about taking part in genetic research may influence insurance companies and/or employers regarding your health.
- Research results obtained in this study will not be placed in your medical record unless we contact you with an important finding and you agree to have it confirmed. We do not think that there will be further risks to your privacy by sharing your samples and/or whole genome information with other researchers; however we cannot predict how genetic information could be used in the future.
- There is a very small risk of bruising or infection from drawing blood similar to what might occur from a routine blood draw that you get for your doctor.

#### 12. If I take part in the Biobank, how will you protect my privacy?

In general, health information that identifies you is private under federal law. However, you should know that in addition to Partners researchers the following people or groups may be able to see, use, and share your identifiable health information from the research and why they may need to do so:
- Any sponsor(s) of this Biobank and the people or groups it hires to help with the Biobank
- The Partners ethics board that oversees the project and the Partners research quality improvement programs
- People from organizations that provide independent accreditation and oversight of hospitals and research
- People or organizations that we hire to do work for us, such as data storage companies, insurers, and lawyers

- Federal and state agencies (such as the Food and Drug Administration, the Department of Health and Human Services, the National Institutes of Health, and other US or foreign government bodies that oversee or review research)
- We share your identifiable health information only when we must, and we ask anyone who receives it from us to protect your privacy. However, once your information is shared outside Partners, we cannot promise that it will remain private.

**13. Certificate of Confidentiality for Health Information and Other Identifying Information from the Research**

To help protect your privacy, we have obtained a Certificate of Confidentiality from the National Institutes of Health. With this Certificate, researchers cannot be forced to disclose information that may identify you, even by a court subpoena, in any federal, state, or local civil, criminal, administrative, legislative, or other proceedings. Researchers will use the Certificate to resist any demands for information that would identify you, except as explained below. The Certificate cannot be used to resist a demand for information from personnel of the United States Government that is used for auditing or evaluation for Federally funded projects or for information that must be disclosed in order to meet the requirements of the Federal Food and Drug Administration (FDA).

You should understand that a Certificate of Confidentiality does not prevent you or a member of your family from voluntarily releasing information about yourself or your involvement in this research. If an insurer, employer, or other person obtains your written consent to receive research information, then the researchers may not use the Certificate to withhold that information.

A Certificate of Confidentiality does not prevent researchers from voluntarily disclosing information about you, without your consent in incidents such as child abuse, and intent to harm yourself or others.

#### Informed Consent and Authorization for Collection of Samples and Health Information for Research

##### Statement of Study Doctor or Person Obtaining Consent

- I have explained the research study to the subject.
- I have answered all questions about this research study to the best of my ability.

______________________________ ___________ ___________
Study Doctor or Person Obtaining Consent Date Time

**Subject Identification**

**Signature of Subject:** I give my consent to take part in this research study and agree to allow my samples and health information to be used and shared as described above.

______________________________ ___________ ___________
Subject Date Time

______________________________
Email

Supplement 2: Letter Informing Participant of Actionable Result

PARTNERS HEALTHCARE | BIOBANK

**Partners HealthCare Biobank**

Dear Mr./Ms. (*Name*),

Thank you for participating in the Partners HealthCare Biobank (Partners Biobank) project at (*Massachusetts General Hospital* OR *Brigham and Women's Hospital*). Researchers using your samples have made discoveries that may be important to your health. A team of experts have reviewed the findings and decided that genetic results from your sample are "actionable", which means that this information would help your doctor make decisions about your care.

This letter is to inform you that you will be contacted by a healthcare professional to discuss your individual research finding. Research findings are not the same as clinical tests done during routine clinical care. Your doctor may want to repeat the research test using a certified clinical laboratory to be certain that the result is correct. The costs for repeating these tests will be billed to your insurance. You can decide whether or not you want the healthcare professional to contact your doctor with your health-related research finding.

If you have any questions about this letter, or about the Partners Biobank study, please contact the Partners Biobank staff by phone at (617-525-6700), or email at. If you would like more information about the Partners Biobank, you may also visit our website at www.partners.org/biobank.

Sincerely,

Scott T. Weiss MD

Scott T. Weiss, MD, MS
PRINCIPAL INVESTIGATOR

### Return of Research Results Phone Scripts

#### Voicemail Message

Hello. This message is for Mr./Ms. (*Name*). My name is [name of genetic counselor] and I am calling from Mass General Brigham.

I am calling with regard to your participation in a research project at (*Massachusetts General Hospital* OR *Brigham and Women's Hospital* OR *Spaulding Rehabilitation Hospital* OR *McLean Hospital*).
Please contact me at [genetic counselor email] or by telephone at [genetic counselor phone].

Have a great day. Goodbye.

### Pre-Test Counseling Phone Call

#### Checklist

| |
|---|
| Name of Subject |
| Subject ID |
| Date of Pre-Test Counseling Phone Call |
| Name of GC |

- ☐ Communicate disease name and description of disease associated with the variant
- ☐ Communicate loss of confidentiality
- ☐ Communicate social and economic disadvantages (impact on insurance)
- ☐ Communicate potential impact on family
- ☐ Communicate cost
    - ☐ Specify that cost of clinical visit to receive result will be billed to patient, but clinical validation will not
- ☐ Review sample collection process: option to do blood draw or saliva kit
    - ☐ If saliva kit, confirm address and phone number to ship kit
    - ☐ If blood draw, schedule visit at CCI
- ☐ Record preference to receive result: apt with MG or HCP
    - ☐ If preference is appointment with MG, we will coordinate with the MG team once the saliva kit has been received by the lab.
    - ☐ If preference is appointment with HCP, ask for the name and contact information for the HCP. This information will be included on the authorization form for the subject to sign. When the patient sends in saliva kit, we will begin to coordinate with the HCP.

PARTNERS HEALTHCARE | BIOBANK

#### Phone Scripts

**GC:**

Hello Mr./Ms. (*Name*), my name is [name of genetic counselor] and I am calling from (*Massachusetts General Hospital* OR *Brigham and Women's Hospital* OR *Spaulding Rehabilitation Hospital* OR *McLean Hospital*).

I am calling with regard to your participation in the Mass General Brigham Biobank project at (*Massachusetts General Hospital* OR *Brigham and Women's Hospital* OR *Spaulding Rehabilitation Hospital* OR *McLean Hospital*). Do you remember joning the Biobank?
If Yes: Procede with below script
If No: The Biobank is a reseach study that you joined to help scientistis learn more about your health. You would have signed a consent form and donated some blood at some point over the last several years.

As part of your consent to participate in the Biobank, we informed you that we would attempt to contact you in the event that research results from your sample might be medically important. I am now calling to let you know that we have a research result from your DNA sample that may be important to your health.

At this point, you have an option to hear more information about this research result, or decline to receive any additional information about the result. If you choose to hear more information about the result, you are still able to decline to receive additional information at any time, for any reason.

Would you like to receive more information about this research result from your DNA sample?

**Subject:**

Yes

**GC:**

The research result we obtained relates to a genetic risk for <insert disease name and description of disease>. This result is NOT a diagnosis, and it does not mean that you will definitely get the disease. At this point, the result only suggests that you may be at risk.

This preliminary research result will require verification in a separate clinical laboratory possibly with a new sample that we can typically get from saliva to confirm that the result is correct. In rare cases, we may need a blood sample if we cannot get enough DNA from the saliva sample. Because this research result was discovered using a non- clinically approved test, it is not recommended that action is taken as a result of this research result at this time, which is why another test must be performed to verify that the result is correct.

Do you feel like you understand what we have discussed so far? Do have any questions at this time?

**Subject:**

Yes/No

*If No, I do not understand:*

**GC:**

Your result was identified as part of a study in a research setting. The standards followed for testing samples may vary among research laboratories. Clinical laboratories are held to higher standards when examining and reporting results and must meet quality standards set by the Clinical Laboratory Improvement Amendments (CLIA). Because of this we must collect a second sample and perform clinical grade testing to be sure the research result we identified is accurate.

*If Yes, I understand:*

**GC:**

Based on this preliminary research result, I recommend that you receive clinical genetic testing to verify the research result.

However, before you make a decision to receive clinical genetic testing, I need to review some important information with you. You will have the option to decline receiving the clinical genetic testing after we review this information.

If the result is clinically confirmed, specific screening, tests, exams, or lifestyle changes may be recommended for early detection and/or disease management. Knowing about a genetic risk variant could also be useful for future financial and/or family planning.

At the moment this variant is a research result and should not be placed in your medical record. However, once you result is clinically confirmed your disclosing provider will note it in medical record and the lab will upload this into your MassGeneral Brigham medical record. This result is also noted in your Mass General Brigham Biobank study record. This means that anyone who has access to your medical record will be able to see your genetic test result, like your other doctors.

As with any information going into your medical record, there is always a potential risk of **loss of confidentiality**. However, we do everything we can to keep your information safe.

Do you feel like you understand the potential loss of confidentiality? Do you have any questions?

**Subject:**

Yes/No

*If yes, GC proceeds to next point.*
*If no, refer to standard questions and answers at the end of this document.*

**GC:**

The second implication concerns potential risk of insurance discrimination. There is a federal law called GINA, which stands for the Genetic Information Non-Discrimination Act, that protects a person from discrimination based on their genetic information in employment and health insurance, but GINA does not provide protection in all situations. In most states, genetic discrimination laws do not currently protect a person from discrimination in life, long-term care, or disability insurance based on genetic information; however, in Massachusetts there are some state laws that offer additional protections. [If asked to elaborate- there must be actuarial evidence to present]. The risks of discrimination are not fully known, but it is possible that an individual known to have a genetic risk factor may have trouble obtaining or keeping life, long-term care, or disability insurance this could potentially cause you **social and economic disadvantages**.

Do you have any questions about the potential risk that the clinical validation could have in terms of access to life, long-term care, or disability insurance?

**Subject:**

Yes/No

*If yes, GC proceeds to next point.*
*If no, refer to standard questions and answers at the end of this document.*

**GC:**

The third implication concerns the potential **impact on your family**. Through clinical genetic testing, you may learn that other family member(s) are at risk for the same condition. Your relatives may or may not want to know this information. [GC to collect additional relevant family history to contextualize the result to their individual family history].

Do you have any questions about the potential impact on your family?

**Subject:**

Yes/No

*If yes, GC proceeds to next point.*
*If no, refer to standard questions and answers at the end of this document.*

**GC:**

The fourth implication is **cost**. There are costs associated with genetic testing. The cost of the sample collection and the test in the clinical laboratory are covered by the Biobank study as long as the test is done at the Laboratory for Molecular Medicine that is affiliated with the Biobank. If the test is done at another laboratory, you and your insurance will need to cover the cost. Once your result is clinically confirmed we recommend you see a doctor to review the implications of your clinical test result and what this means for your health. This doctors visit, typically with a medical geneticist or specialty provider with expertise in genetics, is NOT covered by the study but will be billed to your insurance.

If you or your doctors request follow-up care including medical tests or office visits to follow up on results found in the genetic testing, you may be responsible for these costs; however, these clinical interactions are often covered by health insurance. You or your relatives may be responsible for costs associated with additional testing to learn more about genetic variants that could affect the health of the patient's family.

Do you have any questions about the costs associated with the clinical validation?

**Subject:**

Yes/No

*If yes, GC proceeds to next point.*
*If no, refer to standard questions and answers at the end of this document.*

In order to move forward with the clinical validation, all we would need to do is

*CLIA sample:*
retest a sample of the blood that you already provided to the Biobank. The results of the genetic test should be ready in approximately 3-4 weeks.;

*Non-CLIA sample*:
We will mail you a saliva kit. You will follow the directions included with the tube to collect the saliva and will ship the sample back to us (postage is provided). In very rare cases, we may request that we collect an additional blood sample (if we cannot collect enough DNA though the saliva test).

If you would prefer to provide a blood sample, we can schedule a blood draw with MGH or BWH phlebotomy. In case you chose to do the blood draw, you will need to come into Brigham and Women's Hospital or Mass General Hospital. There will be no cost to you for the blood draw.
The results of the clinical genetic test should be ready in approximately 3-4 weeks.[Note for particularly anxious participants: "We can ask the lab to rush your sample and it should be ready in about 2 weeks, unless they experience lab issues).

Would you like to move forward with the clinical genetic testing?

**Subject:**
Yes/No

**GC:**
*If No:*
*If No:* Thank you for your time. We will send you a survey by mail or email to collect your feedback on our return of research results process. We may also call you to learn more about your decision making and experience. However, if you change your mind, you may contact Mass General Brigham Biobank staff for general questions by phone at (617-525-6700) or email at. We will send you a survey. Goodbye.

----------------------------------------------------------------------------------------------------

*If Yes:*
**GC:**
*CLIA sample*: skip to next section ("Meet with MG/Meet with PCP")

*Non CLIA sample:*

*Saliva sample:*
next step is for me to make sure that we send you a saliva kit. Can you please confirm your address and phone number? We will send the saliva kit by fedex or ups. If you do not receive it in the next few days, please contact the Biobank at 617-525-6700 or email so that we may help you.

*Blood sample:*
OK, the next step is for me to schedule you to come into the Center for Clinical Investigation at Brigham and Women's Hospital or at the Mass General Biobank phlebotomy desk for the clinical blood draw. When would be a good time for you to come in?
<schedule appointment>

Once the test is complete, you will need to meet with a doctor to learn the genetic result. We strongly suggest that you meet with a Medical Geneticist or a specialist with expertise in genetics to review your result and what it means for you and your family. We can help set this up for you. We can also share the result with your HCP if you would like. We will ask you to sign a medical relase form that indicates the doctor(s) you want us to share this information with. We will send this to you with your saliva sample.

If you agree to have your testing done at the MassGeneral Brigham LMM lab, we will also send you a clinical consent form that overviews the fact that you are obtatining a clinical test. We will ask you to sign this and send back with your saliva sample.

[In circumstances where the patient does not want to see genetics, or does not want to come into the city or to a location where there is an available medical genetics provider] If you prefer, you are also welcome to set up an appointment with your Primary Care Physician or other specialty physician. If this is your preference we will ask your permission to speak to your doctor and ask you to sign a medical release so we can share your result with them and we will ask your doctor if he/she is willing to review the results with you. If they do not feel comfortable reviwing these results with you we can help set you up with a specialist. If you agree to have your testing done at the MassGeneral Brigham LMM lab, we will also send you a clinical consent form that overviews the fact that you are obtatining a clinical test. We will ask you to sign this and send back with your saliva sample.

*Would you like to meet with a Medical Geneticist/ Specialist with expertise in genetics [or with your HCP]?*

**Subject:**

Meet with MG/ Specialist vs Meet with HCP

*If meeting with MG, schedule meeting with MG/specialist to be when the results are available. Contact the MG/specialist GC and/or scheduling team to set this up.*

**GC:**

Great. I can schedule a meeting with our Medical Geneticist/specialist for 3-4 weeks from when the lab receives your sample. This will give us enough time to obtain your sample and complete the test.

**GC:**

Great I will send you a medical records release form so that I can talk about and share the test result with your HCP. Once the lab has confirmed your result we will reach out to your HCP to let them know you need to be seen and they should contact you to set up an appointment.

After your results are disclosed you will receive a survey by email or mail asking you questions about participating in the biobank process. We would greatly appreciate you filling out this survey.

**GC:**
Do you have any questions at this time?

**GC:**
*If No:* Alright. Thank you for your time. Goodbye.

*If Yes, GC answers questions or documents questions to discuss in in-person appointment.*

#### Standard Questions and Answers

*Responses to likely questions are as follows. If the participant does not ask these questions over the phone, these will be discussed at the in-person appointment:*

**Subject:** What did you find? Why can't you tell me now?
**GC:** The genetic change/variant identified has been associated with an increased risk for <disease name and description of disease>. Since this research result was not discovered using a clinical genetic test, it is not recommended that any action be taken as a result of this research result at this time. We can assist you in having your variant confirmed in the Mass General Brigham Healthcare clinical laboratory

**Subject:** Does this mean I have a disease? Or will I definitely get the disease?
**GC**: We are returning results to individuals to whom we believe there is may be an increased risk of developing the particular condition. This does not mean you will definitely get the disease. It also depends on other factors such as your age, lifestyle, family history, environment, etc. And we do not know that the research result is correct which is why you would need to have a clinical test to confirm the result.

**Subject:** How can I make a decision about clinical testing if I don't know what you found?
**GC:** The genetic variant identified may be associated with an increased risk for <disease name and description of disease>.  We are informing you about this result since there are guidelines available for screening and ways to lower your risk (if relevant). If you wish to proceed, after the result is confirmed using a clinical genetic test, a medical geneticist/genetic counselor can meet with you and provide you with specific recommendations about your risk, screening and management.

**Subject:** What are the costs of clinical genetic testing?
**GC:**

- Confirming your result using the Mass General Brigham clinical lab would not cost you anything.
- If you or your doctor request medical tests to follow up on results found in the genetic testing, you may be responsible for some of these costs. This may include the cost of additional testing to learn more about genetic variants that could affect the health of your family. To be clear, initial research result confirmation testing will be done at no cost to you, however, if there is follow-up clinical care and additional office visits, costs associated with continued clinical care (e.g. visit co-pays) will be your responsibility.

**Subject:** What are the potential benefits to me?
**GC:**
- Learning about your result may help your doctors make decisions about your medical care; for example, your doctors may recommend specific screening, tests, exams, or lifestyle changes to follow-up on the clinical test result or other conditions. Knowing about a genetic change could also be useful for your future financial and/or family planning and there may be potential benefit to your family members.

**Subject:** What are the potential risks?
**GC:**
- Loss of confidentiality
  Your clinical test result (from the confirmatory testing) will be placed in the electronic medical record as with anything in the medical record there is a potential loss of confidentiality but we do everything we can to keep your information safe and secure. We will not place the research result in the electronic medical record.

- Insurance
  There are laws (GINA) that protect a person from discrimination based on their genetic information in employment and health insurance, but they do not provide protection in all situations. In some states, these laws do not currently protect a person from discrimination in life or disability insurance based on genetic information. The risks of discrimination are not yet known, but it is possible that you may have trouble obtaining or keeping life or disability insurance.

- Impact on families
  You could also learn that other family member(s) are at risk for the same condition. Your relatives may or may not want to know this information.

PARTNERS® HEALTHCARE | BIOBANK

### Survey Reminder Phone Scripts

#### Phone Reminder

Hello Mr./Ms. (*Name*), my name is [name of Biobank staff] and I am calling from (*Massachusetts General Hospital* OR *Brigham and Women's Hospital* OR *Spaulding Rehabilitation Hospital* OR *McLean Hospital*).

I am calling with regard to your participation in the Mass General Brigham Biobank. As part of your consent to participate in this study, you agreed to be sent a few surveys. This is just a reminder to please complete the survey we recently sent you as soon as you can *[For the Baseline survey only- This is just a reminder to please complete the survey prior to your result disclosure visit.]*

Thanks for your time today.

#### Voicemail Message

Hello. This message is for Mr./Ms. (*Name*). My name is [name of Biobank staff] and I am calling from (*Massachusetts General Hospital* OR *Brigham and Women's Hospital* OR *Spaulding Rehabilitation Hospital* OR *McLean Hospital*).

I am calling with regard to your participation in a research project at (*Massachusetts General Hospital* OR *Brigham and Women's Hospital* OR *Spaulding Rehabilitation Hospital* OR *McLean Hospital*).
Please contact me at telephone at [staff phone].

Have a great day. Goodbye.

| Phenotype/Associated Condition | Disease name communicated to patient | Gene | Description of disease communicated to patient |
|---|---|---|---|
| Hereditary Breast and Ovarian Cancer | Hereditary Breast and Ovarian Cancer | BRCA1<br>BRCA2 | A genetic condition that causes an increased risk to develop breast, ovarian, prostate, pancreatic and other cancers. |
| Li-Fraumeni Syndrome | Hereditary Mixed Cancer Syndrome ( different types of cancer) | TP53 | A genetic condition that causes an increased risk to develop several types of cancer, particularly in children and young adults. |
| Peutz-Jeghers Syndrome | Hereditary Gastrointestinal Cancer Syndrome | STK11 | A genetic condition that causes an increased risk to develop breast, ovarian, colorectal, gastric, and pancreatic cancers as well as noncancerous growths, called polyps, in the gastrointestinal tract (stomach and intestines). |
| Lynch Syndrome | Hereditary Colon Cancer Syndrome | MLH1<br>MSH2<br>MSH6<br>PMS2 | A genetic condition that causes an increased risk to develop many types of cancers, particularly colon cancer. In females there is also an increased risk of uterine and ovarian cancer. |
| Familial Adenomatous Polyposis | Hereditary Colon Cancer Syndrome | APC | A genetic condition that causes an increased risk to develop colon cancer. |
| MYH-Associated Polyposis; Adenomas, multiple colorectal, FAP type 2; Colorectal adenomatous polyposis, autosomal recessive, with pilomatricomas | Hereditary Colon Cancer Syndrome | MUTYH | A genetic condition that causes an increased risk to develop colon cancer. |
| Juvenile Polyposis | Hereditary Gastrointestinal Polyp Syndrome | BMRP1<br>SMAD4 | A genetic condition that causes an increased risk to develop cancerous and noncancerous growths, specifically in the gastrointestinal tract (stomach and intestines). |
| Von Hippel Lindau Syndrome | Hereditary Mixed Cancer Syndrome ( different types of cancer) | VHL | A genetic condition that causes an increased risk to develop tumors and cysts in many parts of the body, including the brain, spinal cord, kidneys, and retina. |
| Multiple Endocrine Neoplasia Type 1 | Hereditary Endocrine Cancer Syndrome | MEN1 | A genetic condition that causes an increased risk to develop tumors of the endocrine (hormone-producing) glands including the parathyroid, pancreas, and pituitary gland. |
| Mulitple Endocrine Neoplasia Type 2 | Hereditary Endocrine Cancer Syndrome | RET | A genetic condition that causes an increased risk to develop cancer and non-cancerous tumors of the thyroid and other tumors of hormonal glands. |
| Familial Medullary Thyroid Cancer | Hereditary Thyroid Cancer Syndrome | RET | A genetic condition that causes an increased risk to develop thyroid cancer. |
| PTEN Hamartoma Tumor Syndrome | Hereditary Mixed Cancer Syndrome- sometimes associated with intellectual disabilities | PTEN | A genetic condition that causes an increased risk to develop non-cancerous and cancerous tumors including the thyroid, breast, kidney and uterus. In some individuals this disorder can cause development and learning difficulties. |
| Retinoblastoma | Hereditary Eye Cancer Syndrome | RB1 | A genetic condition that causes increased risk to develop cancer that begins in the eye (retina). There is also an increased risk of other cancers including a gland in the brain (pinealoma), bone, soft tissue, and muscle, and skin cancer. |
| Hereditary Paraganglioma- Pheochromcytoma Syndrome | Hereditary Paraganglioma Syndorme | SDHD<br>SDHAF2<br>SDHC<br>SDHB | A genetic condition that causes increased risk to develop growths on hormone-producing glands that are typically non-cancerous. |
| Tuberous Sclerosis Complex | Hereditary Syndrome that Affects the Brain, Skin and Kidneys | TSC1<br>TSC2 | A genetic condition that causes an increased risk to develop noncancerous tumors in many parts of the body, including the skin, brain, and kidneys. |
| WT1- related Wilms tumor | Hereditary Kidney Cancer Syndrome | WT1 | A genetic condition that causes an increased risk to develop kidney tumors in childhood. |
| Neurofibromatosis type 2 | Hereditary Nervous System Benign Tumor Syndrome | NF2 | A genetic condition that causes an increased risk to develop non-cancerous tumors of the nerves , sometimes causing hearing loss. |

| | | | |
|---|---|---|---|
| Ehlers-Danlos Syndrome- vascular type | Hereditary Connective Tissue Disorder That Can Cause Aortic Dilataion | COL3A1 | A genetic condition that causes an increased risk to develop dilated (enlarged) blood vessels, and rupture of the uterus, intestine and aorta. |
| Marfan Syndrome, Loeys-Dietz Syndromes, and Familial Thoracic Aortic Aneurysms and Dissections | Hereditary Connective Tissue Disorder That Can Cause Aortic Dilataion | FBN1 | A genetic condition that causes an increased risk to develop dilated (enlarged) blood vessels, and rupture of the uterus, intestine and aorta. |
| | | TGFBR1 | |
| | | TGFBR2 | |
| | | SMAD3 | |
| | | ACTA2 | |
| | | MYH11 | |
| Fabry Disease | Hereditary Disorder That Affects The Heart, Kidneys And Some Other Organs | GLA | A genetic condition that causes an increased risk to develop kidney failure, strokes, heart disease, and other medical issues. |
| Hypertrophic cardiomyopathy, Dilated cardiomyopathy | Hereditary Cardiomyopathy (Enlarged Heart) | MYBPC3 | A genetic condition that causes an increased risk to develop a heart problem called cardiomyopathy. This can cause heart failure or an abnormal heart rhythm that can cause the heart to stop beating. |
| | Hereditary Cardiomyopathy (Enlarged Heart) | MYH7 | |
| | Hereditary Cardiomyopathy (Enlarged Heart) | TNNT2 | |
| | Hereditary Cardiomyopathy (Enlarged Heart) | TNNI3 | |
| | Hereditary Cardiomyopathy (Enlarged Heart) | TPM1 | |
| | Hereditary Cardiomyopathy (Enlarged Heart) | MYL3 | |
| | Hereditary Cardiomyopathy (Enlarged Heart) | ACTC1 | |
| | Hereditary Cardiomyopathy (Enlarged Heart) | PRKAG2 | |
| | Hereditary Cardiomyopathy (Enlarged Heart) | MYL2 | |
| | Hereditary Cardiomyopathy (Enlarged Heart) | LMNA | |
| Catecholaminergic polymorphic ventricular tracycardia | Hereditary Heart Rhythm Disorder | RYR2 | A genetic condition that causes an increased risk to develop an abnormal heart rhythm (arrhythmia) that can cause the heart to stop beating. |
| Arrythmogenic right ventricular cadriomyopathy | Hereditary Heart Rhythm Disorder | PKP2 | A genetic condition that causes an increased risk for developing heart problems, such as heart failure or an abnormal heart rhythm that can cause the heart to stop beating. |
| | | DSP | |
| | | DSC2 | |
| | | TMEM43 | |
| | | DSG2 | |
| Romano-Ward Long QT Syndromes Types 1,2, and 3, Brugada Syndrome | Hereditary Heart Rhythm Disorder | KCNQ1 | A genetic condition that causes an increased risk to develop an abnormal heart rhythm (arrhythmia) that can cause the heart to stop beating. |
| | | KCNH2 | |
| | | SCN5A | |
| Familial hypercholesterolemia | Hereditary High Cholesterol Disorder | LDLR | A genetic condition that can cause increased risk to develop high cholesterol levels which can cause problems with the heart and blood vessels. In some people this can be much more significant than typical high cholesterol that many people have. |
| | | APOB | |
| | | PCSK9 | |
| Wilson Disease | Hereditary Copper Metabolism Disorder | ATP7B | A genetic condition that can cause increased risk for copper to build up in the organs which can cause liver failure, mental health issues and abnormal muscle movements. |
| Orinthine transcarbamylase deficiency | Hereditary Urea Cycle Disorder | OTC | A genetic condition that increases the risk for ammonia to accumulate in the blood. This can cause neurological problems including learning issues, confusion and life-threatening comas. |
| Malignant hyperthermia susceptibility | Hereditary Disorder- Causing Abnormal Response to Anesthesia | RYR1 | A genetic condition that increases the risk to develop severe reactions to certain drugs used for general anesthesia. |

Supplement 4: Contacts Needed for Each Participant Outcome

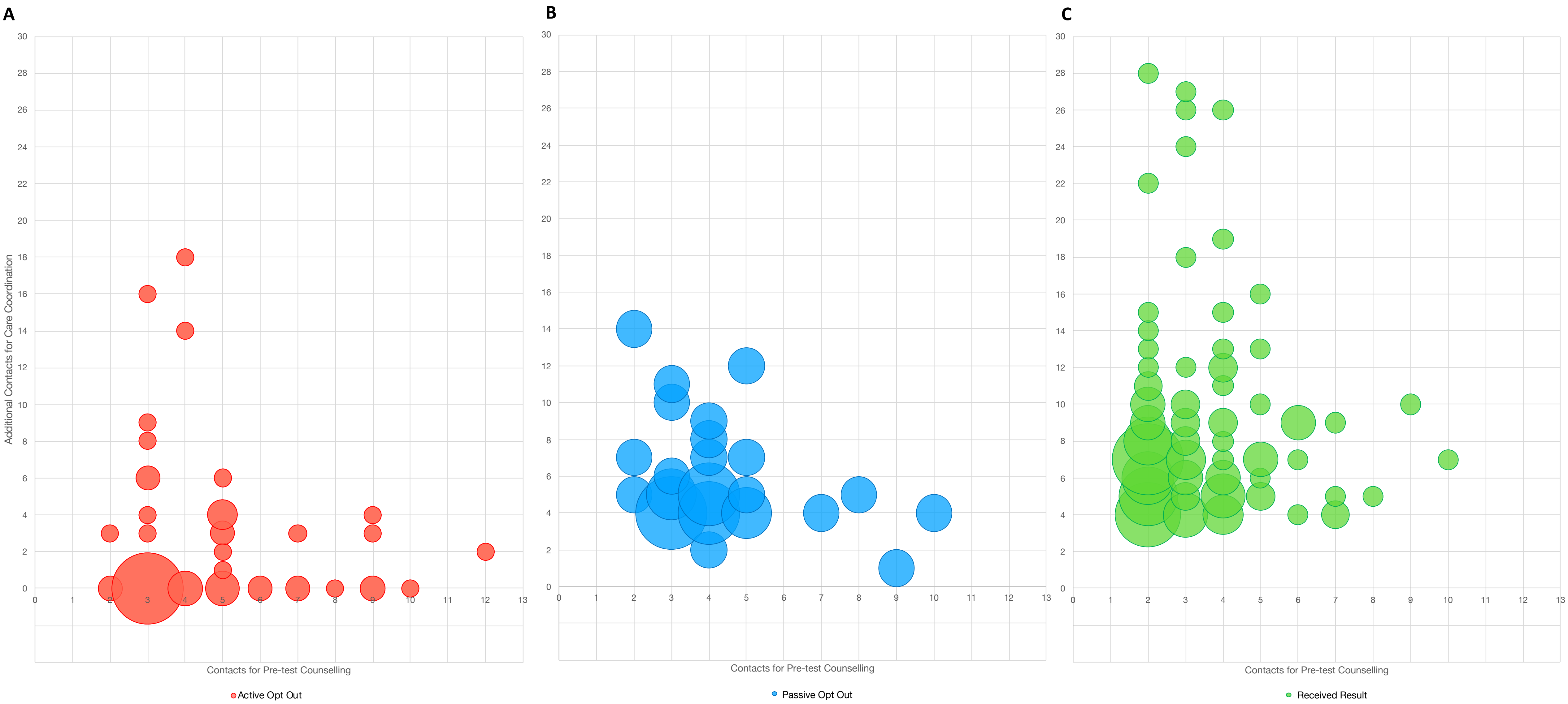

Supplement 5: Example Clinical Report

PARTNERS HEALTHCARE | PERSONALIZED MEDICINE

MASSACHUSETTS GENERAL HOSPITAL    BRIGHAM AND WOMEN'S HOSPITAL

Unit Number(s):

**Laboratory for Molecular Medicine**
65 Landsdowne Street, Cambridge MA 02139
www.partners.org/personalizedmedicine/lmm

Lab Accession:
Patient Name:
Birth Date:
Age Sex:

### MOLECULAR DIAGNOSTICS REPORT

**Specimen Type:**
**Related Accession(s):**
**Referring Physician:**
**Copies To:**

**Received Date:**
**Referring Facility:**
**Referring Fac. MRN:**
**Lab Control Number:**
**Family Number:**

**TEST DESCRIPTION** – Clinical Confirmation Test - Partners Healthcare Biobank RoR
**TEST PERFORMED** – ClinCon-f
**INDICATION FOR TEST** – Partners Biobank Variant Confirmation

#### RESULTS

#### DNA VARIANTS:

Heterozygous c.10580G>A (p.Arg3527Gln), Exon 26, APOB, Pathogenic

#### INTERPRETATION:

**Positive.** DNA sequencing identified the variant listed above. This variant was previously identified by the Partners Biobank research study.

#### INTERPRETATION SUMMARY:

A well-established, heterozygous pathogenic variant in the APOB gene was identified in this individual. Pathogenic variants in APOB are associated with familial hypercholesterolemia (FH); therefore, this individual is at risk for developing hypercholesterolemia. Please note that pathogenic variants in APOB can have reduced penetrance and a less severe phenotype than disease causing LDLR or PCSK9 variants (Youngblom and Knowles, GeneReviews). The available information on this variant is described in the variant interpretation section below.

Disease penetrance and severity can vary due to modifier genes and/or environmental factors. The significance of a variant should therefore be interpreted in the context of the individual's clinical manifestations.

This test is variant specific and does not detect other variants in this gene or other genes.

#### INHERITANCE PATTERN:

FH due to pathogenic variants in the APOB gene is inherited in an autosomal dominant pattern. Each first-degree

#### MOLECULAR DIAGNOSTICS REPORT

relative has a 50% (or 1 in 2) chance of inheriting a variant and its risk for disease. Additional family members may also have inherited this variant and may be at risk for disease.

**VARIANT INTERPRETATION:**

**p.Arg3527Gln, c.10580G>A (APOB; NM_000384.2; Chr2 g.21229160C>T; GRCh37):**

The p.Arg3527Gln variant in APOB is a well-established pathogenic variant that is mainly found in individuals of European descent. It has been previously reported in >500 individuals with familial hypercholesterolemia (FH) and segregated with disease in >50 affected relatives (Soria 1989, März 1993, Leren 1997, Ludwig 1990, Bednarska-Makaruk 2001, Horvath 2001, Kalina 2001). It has also been reported by other clinical laboratories in ClinVar (Variation ID 17890) and has been identified in 53/126056 of European chromosomes, including 1 homozygote, by the Genome Aggregation Database (gnomAD, http://gnomad.broadinstitute.org/; dbSNP rs5742904). This frequency is low enough to be consistent with the frequency of FH in the general population. In summary, this variant meets criteria to be classified as pathogenic for autosomal dominant familial hypercholesterolemia based upon presence in multiple affected individuals and segregation studies. Please note that pathogenic variants in APOB can have reduced penetrance and a less severe phenotype than disease-causing LDLR or PCSK9 variants (Youngblom and Knowles, GeneReviews). ACMG/AMP Criteria applied: PS4_Strong; PP1_Strong.

**RECOMMENDATION:**

Genetic counseling is recommended for this individual and her relatives. Familial variant testing is available for other relatives if desired. For assistance in locating genetic counseling services or disease specialists please call the laboratory at 617-768-8500 or email at. Please note that variant classification, particularly of uncertain significance, may change over time if more information becomes available. Please contact us at 617-7688500 or.

**COMMENTS:**

An online research opportunity called GenomeConnect is available for any recipient of genetic testing to advance knowledge of genetic variants by sharing de-identified genetic and health information. Please visit genomeconnect.org to learn more.

**TEST INFORMATION BACKGROUND:**

Clinical confirmation testing is performed to assess an individual's risk for developing or being a carrier of a genetic condition, determine if a variant has occurred de novo, determine cis or trans configuration, or to help clarify the significance of a variant through segregation studies. An individual's risk of disease depends on several factors, including the interpretation of the variant, penetrance and inheritance of the disease, presence of other variants, as well as other genetic and environmental factors.

**METHODOLOGY:**

Clinical confirmation testing is performed by Sanger sequencing of the variant(s) in the patient. This method is over 99.9% accurate. Testing may include specialized methods to address technically difficult genomic regions. Variant classifications are based on ACMG/AMP criteria (Richards et al. 2015) with ClinGen rule specifications

#### MOLECULAR DIAGNOSTICS REPORT

(https://www.clinicalgenome.org/working-groups/sequence-variant-interpretation/). Variants are reported according to HGVS nomenclature (www.hgvs.org/mutnomen). This test was developed and its performance characteristics determined by the Laboratory for Molecular Medicine at Partners Healthcare Personalized Medicine (LMM, 65 Landsdowne St, Cambridge, MA 02139; 617-768-8500; CLIA#22D1005307). It has not been cleared or approved by the U.S. Food and Drug Administration (FDA). The FDA has determined that such clearance or approval is not necessary.

REPORT PREPARATION by
REPORT by
Final Diagnosis by

#### Biobank ROR Decliner Interview Guide

Hello, Mr./Mrs. ________. My name is XX and I am calling from the Partners Biobank at Brigham and Women's Hospital. You were previously contacted by the biobank in (month, year) about your research genetic test results. At that time, you did not want to pursue this information further and have these results confirmed by a clinical laboratory. The biobank staff supports any decision that participants make, and I am reaching out to you today to try to better understand how participants make these decisions about receiving or not receiving their genetic results. If you are willing, I would like to interview you for about 20-30 minutes.

**Would you be willing to participate?**

**If they respond yes:**

The purpose of this interview is to hear your thoughts about your decision to not receive your genetic test results. There are no right or wrong answers to these questions. We just want to better understand what our biobank participants think about when faced with this decision. This interview will be audio recorded and then later written out on paper to be analyzed. All identifying information will be removed in the paper version and all of the analyzed data will remain anonymous. I am going to begin the recording now.

This interview will last about 20-30 minutes. For participating in this interview, we will send you a $15 Amazon gift card as a thank you for your time. Your participation in this interview is completely voluntary and if at any point you wish to stop the interview, you are welcome to do so. Additionally, if there is a question you would rather not answer, just let me know.

Do you have any questions before we get started?

1. Can you tell me a little bit about the reasons why you declined to receive your genetic result?
   - **If a participant endorses one of the reasons below, can consider the prompt questions**
   - Did not want to provide another sample
   - Logistics/ Time Commitment
   - Anxiety of receiving results
   - Anxiety of waiting for results (**prompt:** Did you feel that waiting for the result would lead to more anxiety?)
   - Overwhelmed (not a good time) (**prompt:** Were there too many other things that were going on at the time that you were initially contacted?)
   - Simply not interested

- Concerned about confidentiality of test results (**prompt:** Could you say a little more about these concerns? Who were you worried would learn the information? Why were you worried?**)**
- Concerned about potential to receive unfavorable results (**prompt:** Could you say a little more about these concerns?)
- Concerned about genetics test report in medical record (**prompt:** If you could have chosen whether or not your genetic test result would be put into your medical record, would you have decided to receive your genetic result? )
- Religious or spiritual reasons (**prompt:** Did your religious or spiritual believed lead you to decline receiving your genetic results?
- Concerned about insurance discrimination (**prompt:** Were you worried about your genetic results impacting your insurance coverage?)
- Other (**prompt:** Were there any other factors that you considered when deciding not to receive your genetic test results?)

2. **If they endorsed "concerned about confidentiality of test results":** If you could have chosen whether or not your genetic test results would be put into your medical record, would you have decided to receive your genetic result?

3. Please share with me what you remember about donating a blood sample to the Partners biobank?
    - Can you tell me a little bit about why you decided to donate a sample?
    - Did you know that genetic testing might be performed on your sample?
        i. **If yes:** Did that knowledge influence your decision to provide a sample to the biobank?
        ii. **If no:** What was the main reason you decided to join the biobank?
    - **If they do not remember providing a sample:** You enrolled in the biobank during a visit to (Clinic they were enrolled at) and donated a sample for research, does that appointment help you recall joining?

4. (If participant recalls joining) When you enrolled in the biobank, what were your expectations in terms of hearing back about any results they might find?
    - (For both people who recall enrolling and those who don't): what was your initial reaction to being contact by the Biobank about a potentially actionable result related to the sample you provided?

5. To remind you, someone with the biobank contacted you about genetic testing results. Do you remember what condition these results were related to?
    - If **yes:** How familiar are you with this condition?
    - If **No:** The biobank contacted, you with genetic test results related to ____ condition. How familiar are you with this condition?
    - Can you tell me a little bit about the experiences you have had with this condition?
        i. Do you yourself have any symptoms of this condition like (describe symptoms specific to the condition)
        ii. Do you have any family members with this condition? If so, who?

iii. Do you know of anyone else in your life with this condition?
iv. What are your general thoughts about the condition?

6. When you spoke to the biobank genetic counselor, you told her were not interested in having your genetic result confirmed by a clinical laboratory. Do you remember making that decision?
   - Did any of these experiences or opinions I noted above impact your decision to decline to have your research result confirmed?
   - Can you tell me about your reaction to receive that call?
   - What did you think about the information the genetic counselor provided?
   - Can you tell me a little more about why you decide to opt out of having your research result confirmed?
     i. What were the main factors that you thought about when making that decision?
        1. What other things were going on at the time that played a role in your decision?
        2. How did the thought of how your result might impact family affect your decision?
        3. Were you worried about who would have access to this genetic test report?
           a. If yes were there specific people or organizations you were worried might gain access?
              i. Healthcare providers
              ii. Insurers (if yes to this ask them to elaborate)
              iii. Other
     ii. Did you consult with anyone when making this decision?
        1. Spouse, parent, friend, sibling, other family member?
     iii. Some individuals are able to make decisions very quickly while others take a little bit more time to come to a decision. Where would you say you fall on this spectrum? Do you remember how long it took you to make this decision about your Biobank result?
     iv. How difficult was it to decide whether or not to receive your genetic test result?

7. Can you tell me about the experience you have had with genetics and/or genomic sequencing outside of the Partners biobank project?
   - Educational experiences, personal experiences, professional experiences?
   - Did these experiences influence your decision about your research result?

8. Is there anything else you'd like to tell me about your experience that we haven't covered?
   - Do you have any advice for others participating in a biobank?

**Conclusion:** Thank you so much for taking the time out of your day to complete this interview, I greatly appreciate it. We have this address on file for you, would this be the best place to send your Amazon gift card? (read address we have on file).

[If the participant expressed interest in learning their genetic testing result, or has new concerns, I will provide the contact information for the Biobank counselor who can talk about their result in more detail and discuss next steps.]

Thank you again for your time, I hope you have a great rest of your day!

**If participants do not answer the phone call, the following voicemail will be left:**

Hello. This message is for Mr./Mrs. (*Name).* My name is XXX and I am calling from Brigham and Women's Hospital in regard to your participation in the Partners Healthcare Biobank. Please contact me by telephone at 617-529-7738.

Thank you and have a great day. Goodbye.

*We will also interview participants who initially opted to have their result confirmed but later opted out directly or passively by not sending their saliva kit back in or not coming for their clinical appointment, after reminders from the study team.

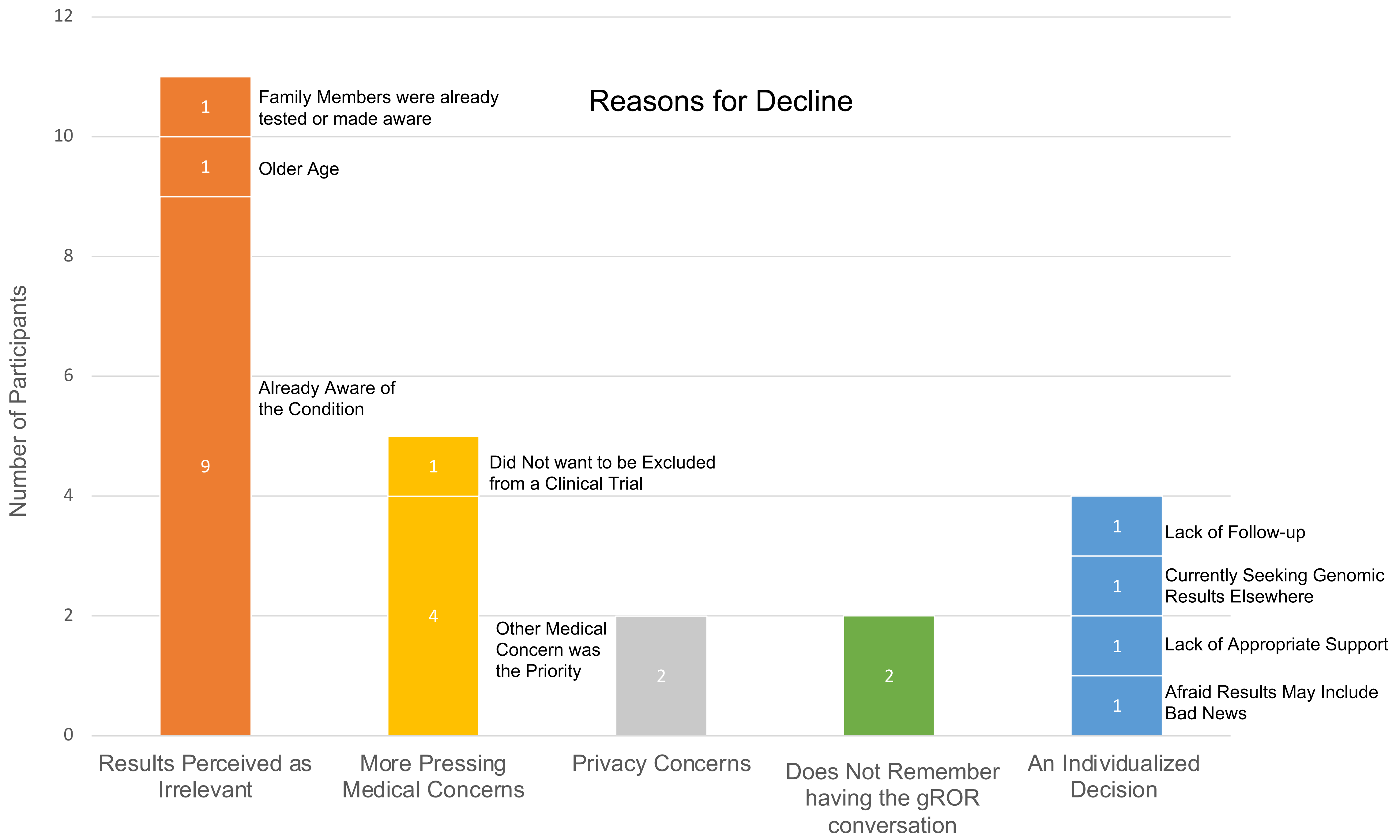
Reasons for Decline
Number of Participants
12
10
8
6
4
2
0
1
Family Members were already tested or made aware
1
Older Age
9
Already Aware of the Condition
1
Did Not want to be Excluded from a Clinical Trial
4
Other Medical Concern was the Priority
2
2
1
Lack of Follow-up
1
Currently Seeking Genomic Results Elsewhere
1
Lack of Appropriate Support
1
Afraid Results May Include Bad News
Results Perceived as Irrelevant
More Pressing Medical Concerns
Privacy Concerns
Does Not Remember having the gROR conversation
An Individualized Decision
