## Supplementary material for "Returning Actionable Genomic Results in a Research Biobank: Analytic Validity, Clinical Implementation and Resource Utilization": COI disclosure forms

### ICMJE DISCLOSURE FORM

**Date:_8/2/21____________________________________________________________________________**
**Your Name:_Carrie L. Blout Zawatsky_______________________________________________________**
**Manuscript Title:_** Returning Actionable Genomic Results in a Research Biobank: Analytic Validity, Clinical Implementation and Resource Utilization
**___________________________________________________________________________**
**Manuscript number (if known):_______________________________________________________**

**In the interest of transparency, we ask you to disclose all relationships/activities/interests listed below that are related to the content of your manuscript. "Related" means any relation with for-profit or not-for-profit third parties whose interests may be affected by the content of the manuscript. Disclosure represents a commitment to transparency and does not necessarily indicate a bias. If you are in doubt about whether to list a relationship/activity/interest, it is preferable that you do so.**

**The following questions apply to the author's relationships/activities/interests as they relate to the current manuscript only.**

**The author's relationships/activities/interests should be defined broadly. For example, if your manuscript pertains to the epidemiology of hypertension, you should declare all relationships with manufacturers of antihypertensive medication, even if that medication is not mentioned in the manuscript.**

**In item #1 below, report all support for the work reported in this manuscript without time limit. For all other items, the time frame for disclosure is the past 36 months.**

| | | **Name all entities with whom you have this relationship or indicate none (add rows as needed)** | **Specifications/Comments (e.g., if payments were made to you or to your institution)** |
|---|---|---|---|
| | | **Time frame: Since the initial planning of the work** | |
| 1 | All support for the present manuscript (e.g., funding, provision of study materials, medical writing, article processing charges, etc.) **No time limit for this item.** | ____ None<br>NIH HG008685 | |
| | | **Time frame: past 36 months** | |
| 2 | Grants or contracts from any entity (if not indicated in item #1 above). | _X___ None | |
| 3 | Royalties or licenses | __X__ None | |

| | | |
|---|---|---|
| 4 | Consulting fees | __X__ None |
| 5 | Payment or honoraria for lectures, presentations, speakers bureaus, manuscript writing or educational events | __X__ None |
| 6 | Payment for expert testimony | __X__ None |
| 7 | Support for attending meetings and/or travel | __X__ None |
| 8 | Patents planned, issued or pending | __X__ None |
| 9 | Participation on a Data Safety Monitoring Board or Advisory Board | __X__ None |
| 10 | Leadership or fiduciary role in other board, society, committee or advocacy group, paid or unpaid | _X___ None |
| 11 | Stock or stock options | __X__ None |
| 12 | Receipt of equipment, materials, drugs, medical writing, gifts or other services | _X___ None |
| 13 | Other financial or non-financial interests | __X__ None |

**Please place an "X" next to the following statement to indicate your agreement:**

**_X__ I certify that I have answered every question and have not altered the wording of any of the questions on this form.**

**ICMJE DISCLOSURE FORM**

**Date: 7/29/2021**
**Your Name: Natalie Boutin**
**Manuscript Title:_** Returning Actionable Genomic Results in a Research Biobank: Analytic Validity, Clinical Implementation and Resource Utilization
______________________________________________________________________
**Manuscript number (if known):___________________________________________________________________**

**In the interest of transparency, we ask you to disclose all relationships/activities/interests listed below that are related to the content of your manuscript. "Related" means any relation with for-profit or not-for-profit third parties whose interests may be affected by the content of the manuscript. Disclosure represents a commitment to transparency and does not necessarily indicate a bias. If you are in doubt about whether to list a relationship/activity/interest, it is preferable that you do so.**

**The following questions apply to the author's relationships/activities/interests as they relate to the current manuscript only.**

**The author's relationships/activities/interests should be defined broadly. For example, if your manuscript pertains to the epidemiology of hypertension, you should declare all relationships with manufacturers of antihypertensive medication, even if that medication is not mentioned in the manuscript.**

**In item #1 below, report all support for the work reported in this manuscript without time limit. For all other items, the time frame for disclosure is the past 36 months.**

| | | **Name all entities with whom you have this relationship or indicate none (add rows as needed)** | **Specifications/Comments (e.g., if payments were made to you or to your institution)** |
|---|---|---|---|
| | | **Time frame: Since the initial planning of the work** | |
| 1 | All support for the present manuscript (e.g., funding, provision of study materials, medical writing, article processing charges, etc.) **No time limit for this item.** | __x__ None | |
| | | **Time frame: past 36 months** | |
| 2 | Grants or contracts from any entity (if not indicated in item #1 above). | __x__ None | |
| 3 | Royalties or licenses | __x__ None | |

| | | |
|---|---|---|
| 4 | Consulting fees | __x__ None |
| 5 | Payment or honoraria for lectures, presentations, speakers bureaus, manuscript writing or educational events | __x__ None |
| 6 | Payment for expert testimony | __x__ None |
| 7 | Support for attending meetings and/or travel | __x__ None |
| 8 | Patents planned, issued or pending | __x__ None |
| 9 | Participation on a Data Safety Monitoring Board or Advisory Board | __x__ None |
| 10 | Leadership or fiduciary role in other board, society, committee or advocacy group, paid or unpaid | __x__ None |
| 11 | Stock or stock options | __x__ None |
| 12 | Receipt of equipment, materials, drugs, medical writing, gifts or other services | ___x_ None |
| 13 | Other financial or non-financial interests | __x__ None |

**Please place an "X" next to the following statement to indicate your agreement:**

**_x__ I certify that I have answered every question and have not altered the wording of any of the questions on this form.**

#### ICMJE DISCLOSURE FORM

**Date:**______________**7/28/21**__________________________________________________________________________
**Your Name:**__**Kurt Christensen**________________________________________________________________________
**Manuscript Title:**_ Returning Actionable Genomic Results in a Research Biobank: Analytic Validity, Clinical Implementation and Resource Utilization
___________________________________________________________________________
**Manuscript number (if known):**_________________________________________________

**In the interest of transparency, we ask you to disclose all relationships/activities/interests listed below that are related to the content of your manuscript. "Related" means any relation with for-profit or not-for-profit third parties whose interests may be affected by the content of the manuscript. Disclosure represents a commitment to transparency and does not necessarily indicate a bias. If you are in doubt about whether to list a relationship/activity/interest, it is preferable that you do so.**

**The following questions apply to the author's relationships/activities/interests as they relate to the current manuscript only.**

**The author's relationships/activities/interests should be defined broadly. For example, if your manuscript pertains to the epidemiology of hypertension, you should declare all relationships with manufacturers of antihypertensive medication, even if that medication is not mentioned in the manuscript.**

**In item #1 below, report all support for the work reported in this manuscript without time limit. For all other items, the time frame for disclosure is the past 36 months.**

| | | **Name all entities with whom you have this relationship or indicate none (add rows as needed)** | **Specifications/Comments (e.g., if payments were made to you or to your institution)** |
|---|---|---|---|
| | | **Time frame: Since the initial planning of the work** | |
| 1 | All support for the present manuscript (e.g., funding, provision of study materials, medical writing, article processing charges, etc.) **No time limit for this item.** | __X__ None | |
| | | **Time frame: past 36 months** | |
| 2 | Grants or contracts from any entity (if not indicated in item #1 above). | Grants from NIH and Sanford Health | Payments to Harvard Pilgrim Health Care Institute |
| 3 | Royalties or licenses | ____ UpToDate | Payments from educational articles about genomic secondary findings |

| | | |
|---|---|---|
| 4 | Consulting fees | __X__ None |
| 5 | Payment or honoraria for lectures, presentations, speakers bureaus, manuscript writing or educational events | __X__ None |
| 6 | Payment for expert testimony | _X___ None |
| 7 | Support for attending meetings and/or travel | __X__ None |
| 8 | Patents planned, issued or pending | __X__ None |
| 9 | Participation on a Data Safety Monitoring Board or Advisory Board | __X__ None |
| 10 | Leadership or fiduciary role in other board, society, committee or advocacy group, paid or unpaid | __X__ None |
| 11 | Stock or stock options | __X__ None |
| 12 | Receipt of equipment, materials, drugs, medical writing, gifts or other services | __X__ None |
| 13 | Other financial or non-financial interests | __X__ None |

**Please place an "X" next to the following statement to indicate your agreement:**

**_X__ I certify that I have answered every question and have not altered the wording of any of the questions on this form.**

### ICMJE DISCLOSURE FORM

**Date:___7/29/21______________________________________________________________________**
**Your Name:___EmmaPerez____________________________________________________________________**
**Manuscript Title:_** Returning Actionable Genomic Results in a Research Biobank: Analytic Validity, Clinical Implementation and Resource Utilization
**___________________________________________________________________________**
**Manuscript number (if known):_________________________________________________________________**

**In the interest of transparency, we ask you to disclose all relationships/activities/interests listed below that are related to the content of your manuscript. "Related" means any relation with for-profit or not-for-profit third parties whose interests may be affected by the content of the manuscript. Disclosure represents a commitment to transparency and does not necessarily indicate a bias. If you are in doubt about whether to list a relationship/activity/interest, it is preferable that you do so.**

**The following questions apply to the author's relationships/activities/interests as they relate to the current manuscript only.**

**The author's relationships/activities/interests should be defined broadly. For example, if your manuscript pertains to the epidemiology of hypertension, you should declare all relationships with manufacturers of antihypertensive medication, even if that medication is not mentioned in the manuscript.**

**In item #1 below, report all support for the work reported in this manuscript without time limit. For all other items, the time frame for disclosure is the past 36 months.**

| | | **Name all entities with whom you have this relationship or indicate none (add rows as needed)** | **Specifications/Comments (e.g., if payments were made to you or to your institution)** |
|---|---|---|---|
| | | **Time frame: Since the initial planning of the work** | |
| 1 | All support for the present manuscript (e.g., funding, provision of study materials, medical writing, article processing charges, etc.) **No time limit for this item.** | NIH - HG008685 | Partial salary from Mass General Brigham eMERGE III grant |
| | | **Time frame: past 36 months** | |
| 2 | Grants or contracts from any entity (if not indicated in item #1 above). | X None | |
| 3 | Royalties or licenses | X None | |

| | | | |
|---|---|---|---|
| 4 | Consulting fees | X None | |
| 5 | Payment or honoraria for lectures, presentations, speakers bureaus, manuscript writing or educational events | ___ None | |
| | | MGH Institute of Health Professions | Lecture honoraria. Some content from manuscript included in lecture. |
| 6 | Payment for expert testimony | X None | |
| 7 | Support for attending meetings and/or travel | ___ None | |
| | | National Society of Genetic Counselors Conference 2020 | Presented poster using data from manuscript |
| | | eMERGE conferences | Funded by NIH - HG008685 |
| 8 | Patents planned, issued or pending | X None | |
| 9 | Participation on a Data Safety Monitoring Board or Advisory Board | X None | |
| 10 | Leadership or fiduciary role in other board, society, committee or advocacy group, paid or unpaid | X None | |
| 11 | Stock or stock options | X None | |
| 12 | Receipt of equipment, materials, drugs, medical writing, gifts or other services | X None | |
| 13 | Other financial or non-financial interests | X None | |

**Please place an "X" next to the following statement to indicate your agreement:**

**X I certify that I have answered every question and have not altered the wording of any of the questions on this form.**

#### ICMJE DISCLOSURE FORM

**Date:**___7/30/2021_______________________________________________________________________
**Your Name:**___Elizabeth Karlson, MD.____________________________________________________________
**Manuscript Title:**_ Returning Actionable Genomic Results in a Research Biobank: Analytic Validity, Clinical Implementation and Resource Utilization
_________________________________________________________________________________
**Manuscript number (if known):**______________________________________________________________

**In the interest of transparency, we ask you to disclose all relationships/activities/interests listed below that are related to the content of your manuscript. "Related" means any relation with for-profit or not-for-profit third parties whose interests may be affected by the content of the manuscript. Disclosure represents a commitment to transparency and does not necessarily indicate a bias. If you are in doubt about whether to list a relationship/activity/interest, it is preferable that you do so.**

**The following questions apply to the author's relationships/activities/interests as they relate to the current manuscript only.**

**The author's relationships/activities/interests should be defined broadly. For example, if your manuscript pertains to the epidemiology of hypertension, you should declare all relationships with manufacturers of antihypertensive medication, even if that medication is not mentioned in the manuscript.**

**In item #1 below, report all support for the work reported in this manuscript without time limit. For all other items, the time frame for disclosure is the past 36 months.**

| | | **Name all entities with whom you have this relationship or indicate none (add rows as needed)** | **Specifications/Comments (e.g., if payments were made to you or to your institution)** |
|---|---|---|---|
| | | **Time frame: Since the initial planning of the work** | |
| 1 | All support for the present manuscript (e.g., funding, provision of study materials, medical writing, article processing charges, etc.) **No time limit for this item.** | ____ None | |
| | | NIH U01HG008685 | |
| | | **Time frame: past 36 months** | |
| 2 | Grants or contracts from any entity (if not indicated in item #1 above). | NIH 1OT2OD026553 | |
| 3 | Royalties or licenses | __X__ None | |

| | | |
|---|---|---|
| 4 | Consulting fees | __X__ None |
| 5 | Payment or honoraria for lectures, presentations, speakers bureaus, manuscript writing or educational events | __X__ None |
| 6 | Payment for expert testimony | _X___ None |
| 7 | Support for attending meetings and/or travel | __X__ None |
| 8 | Patents planned, issued or pending | __X__ None |
| 9 | Participation on a Data Safety Monitoring Board or Advisory Board | __X__ None |
| 10 | Leadership or fiduciary role in other board, society, committee or advocacy group, paid or unpaid | __X__ None |
| 11 | Stock or stock options | __X__ None |
| 12 | Receipt of equipment, materials, drugs, medical writing, gifts or other services | __X__ None |
| 13 | Other financial or non-financial interests | __X__ None |

**Please place an "X" next to the following statement to indicate your agreement:**

**__X_ I certify that I have answered every question and have not altered the wording of any of the questions on this form.**

#### ICMJE DISCLOSURE FORM

**Date:_July 27, 2021______________________________________________________________________________**
**Your Name:__Nina B. Gold, MD__________________________________________________________________**
**Manuscript Title:_** Returning Actionable Genomic Results in a Research Biobank: Analytic Validity, Clinical Implementation and Resource Utilization
__________________________________________________________________________
**Manuscript number (if known):_______________________________________________________________**

**In the interest of transparency, we ask you to disclose all relationships/activities/interests listed below that are related to the content of your manuscript. "Related" means any relation with for-profit or not-for-profit third parties whose interests may be affected by the content of the manuscript. Disclosure represents a commitment to transparency and does not necessarily indicate a bias. If you are in doubt about whether to list a relationship/activity/interest, it is preferable that you do so.**

**The following questions apply to the author's relationships/activities/interests as they relate to the current manuscript only.**

**The author's relationships/activities/interests should be defined broadly. For example, if your manuscript pertains to the epidemiology of hypertension, you should declare all relationships with manufacturers of antihypertensive medication, even if that medication is not mentioned in the manuscript.**

**In item #1 below, report all support for the work reported in this manuscript without time limit. For all other items, the time frame for disclosure is the past 36 months.**

| | | **Name all entities with whom you have this relationship or indicate none (add rows as needed)** | **Specifications/Comments (e.g., if payments were made to you or to your institution)** |
|---|---|---|---|
| | | **Time frame: Since the initial planning of the work** | |
| 1 | All support for the present manuscript (e.g., funding, provision of study materials, medical writing, article processing charges, etc.) **No time limit for this item.** | __x__ None | |
| | | **Time frame: past 36 months** | |
| 2 | Grants or contracts from any entity (if not indicated in item #1 above). | ____ None | Eleanor and Miles Shore Faculty Development Award at Massachusetts General Hospital<br>Greenwall Foundation grant |
| 3 | Royalties or licenses | __x__ None | |

| | | | |
|---|---|---|---|
| 4 | Consulting fees | __x__ None | |
| 5 | Payment or honoraria for lectures, presentations, speakers bureaus, manuscript writing or educational events | __x__ None | |
| 6 | Payment for expert testimony | __x__ None | |
| 7 | Support for attending meetings and/or travel | ____ None | Eleanor and Miles Shore Faculty Development Award at Massachusetts General Hospital |
| 8 | Patents planned, issued or pending | __x__ None | |
| 9 | Participation on a Data Safety Monitoring Board or Advisory Board | __x__ None | |
| 10 | Leadership or fiduciary role in other board, society, committee or advocacy group, paid or unpaid | ____ None | American College of Medical Genetics, Practice Guideline and Policy Committee member |
| 11 | Stock or stock options | __x__ None | |
| 12 | Receipt of equipment, materials, drugs, medical writing, gifts or other services | __x__ None | |
| 13 | Other financial or non-financial interests | __x__ None | |

**Please place an "X" next to the following statement to indicate your agreement:**

**_x__ I certify that I have answered every question and have not altered the wording of any of the questions on this form.**

**ICMJE DISCLOSURE FORM**

**Date:**__7/29/21________________________________________________________________________________
**Your Name:**__Hana Zouk________________________________________________________________________________
**Manuscript Title:**_ Returning Actionable Genomic Results in a Research Biobank: Analytic Validity, Clinical Implementation and Resource Utilization
________________________________________________________________________________
**Manuscript number (if known):**________________________________________________________________________

**In the interest of transparency, we ask you to disclose all relationships/activities/interests listed below that are related to the content of your manuscript. "Related" means any relation with for-profit or not-for-profit third parties whose interests may be affected by the content of the manuscript. Disclosure represents a commitment to transparency and does not necessarily indicate a bias. If you are in doubt about whether to list a relationship/activity/interest, it is preferable that you do so.**

**The following questions apply to the author's relationships/activities/interests as they relate to the current manuscript only.**

**The author's relationships/activities/interests should be defined broadly. For example, if your manuscript pertains to the epidemiology of hypertension, you should declare all relationships with manufacturers of antihypertensive medication, even if that medication is not mentioned in the manuscript.**

**In item #1 below, report all support for the work reported in this manuscript without time limit. For all other items, the time frame for disclosure is the past 36 months.**

| | | **Name all entities with whom you have this relationship or indicate none (add rows as needed)** | **Specifications/Comments (e.g., if payments were made to you or to your institution)** |
|---|---|---|---|
| **Time frame: Since the initial planning of the work** | | | |
| 1 | All support for the present manuscript (e.g., funding, provision of study materials, medical writing, article processing charges, etc.) **No time limit for this item.** | ____ None | |
| | | National Human Genome Research Institute (NHGRI) funding: U01HG008685 & U01HG008676 (Role: Co-Investigator) | |
| **Time frame: past 36 months** | | | |
| 2 | Grants or contracts from any entity (if not indicated in item #1 above). | ___X_ None | |
| 3 | Royalties or licenses | _X___ None | |

| | | |
|---|---|---|
| 4 | Consulting fees | _X___ None |
| 5 | Payment or honoraria for lectures, presentations, speakers bureaus, manuscript writing or educational events | _X___ None |
| 6 | Payment for expert testimony | _X___ None |
| 7 | Support for attending meetings and/or travel | __ X__ None |
| 8 | Patents planned, issued or pending | _X___ None |
| 9 | Participation on a Data Safety Monitoring Board or Advisory Board | __X__ None |
| 10 | Leadership or fiduciary role in other board, society, committee or advocacy group, paid or unpaid | _X___ None |
| 11 | Stock or stock options | ___X_ None |
| 12 | Receipt of equipment, materials, drugs, medical writing, gifts or other services | __X__ None |
| 13 | Other financial or non-financial interests | ___X_ None |

**Please place an "X" next to the following statement to indicate your agreement:**

**_X__ I certify that I have answered every question and have not altered the wording of any of the questions on this form.**

#### ICMJE DISCLOSURE FORM

**Date:** **7/29/2021**______________________________________________

**Your Name:** **Kalotina Machini**______________________________________________

**Manuscript Title:_** Returning Actionable Genomic Results in a Research Biobank: Analytic Validity, Clinical Implementation and Resource Utilization

______________________________________________

**Manuscript number (if known):**______________________________________________

**In the interest of transparency, we ask you to disclose all relationships/activities/interests listed below that are related to the content of your manuscript. "Related" means any relation with for-profit or not-for-profit third parties whose interests may be affected by the content of the manuscript. Disclosure represents a commitment to transparency and does not necessarily indicate a bias. If you are in doubt about whether to list a relationship/activity/interest, it is preferable that you do so.**

**The following questions apply to the author's relationships/activities/interests as they relate to the current manuscript only.**

**The author's relationships/activities/interests should be defined broadly. For example, if your manuscript pertains to the epidemiology of hypertension, you should declare all relationships with manufacturers of antihypertensive medication, even if that medication is not mentioned in the manuscript.**

**In item #1 below, report all support for the work reported in this manuscript without time limit. For all other items, the time frame for disclosure is the past 36 months.**

| | | **Name all entities with whom you have this relationship or indicate none (add rows as needed)** | **Specifications/Comments (e.g., if payments were made to you or to your institution)** |
|---|---|---|---|
| | | **Time frame: Since the initial planning of the work** | |
| 1 | All support for the present manuscript (e.g., funding, provision of study materials, medical writing, article processing charges, etc.) **No time limit for this item.** | _x___ None | |
| | | **Time frame: past 36 months** | |
| 2 | Grants or contracts from any entity (if not indicated in item #1 above). | __x__ None | |
| 3 | Royalties or licenses | __x__ None | |

| | | |
|---|---|---|
| 4 | Consulting fees | __x__ None |
| 5 | Payment or honoraria for lectures, presentations, speakers bureaus, manuscript writing or educational events | _x__ None |
| 6 | Payment for expert testimony | __x__ None |
| 7 | Support for attending meetings and/or travel | ___x_ None |
| 8 | Patents planned, issued or pending | __x__ None |
| 9 | Participation on a Data Safety Monitoring Board or Advisory Board | __x__ None |
| 10 | Leadership or fiduciary role in other board, society, committee or advocacy group, paid or unpaid | __x__ None |
| 11 | Stock or stock options | __x__ None |
| 12 | Receipt of equipment, materials, drugs, medical writing, gifts or other services | __x__ None |
| 13 | Other financial or non-financial interests | __x__ None |

**Please place an "X" next to the following statement to indicate your agreement:**

**_X__ I certify that I have answered every question and have not altered the wording of any of the questions on this form.**

#### ICMJE DISCLOSURE FORM

**Date: 7/28/2021**
**Your Name: Christopher Koch**
**Manuscript Title:_** Returning Actionable Genomic Results in a Research Biobank: Analytic Validity, Clinical Implementation and Resource Utilization
______________________________________________________________________
**Manuscript number (if known):____________________________________________________________**

**In the interest of transparency, we ask you to disclose all relationships/activities/interests listed below that are related to the content of your manuscript. "Related" means any relation with for-profit or not-for-profit third parties whose interests may be affected by the content of the manuscript. Disclosure represents a commitment to transparency and does not necessarily indicate a bias. If you are in doubt about whether to list a relationship/activity/interest, it is preferable that you do so.**

**The following questions apply to the author's relationships/activities/interests as they relate to the current manuscript only.**

**The author's relationships/activities/interests should be defined broadly. For example, if your manuscript pertains to the epidemiology of hypertension, you should declare all relationships with manufacturers of antihypertensive medication, even if that medication is not mentioned in the manuscript.**

**In item #1 below, report all support for the work reported in this manuscript without time limit. For all other items, the time frame for disclosure is the past 36 months.**

| | | **Name all entities with whom you have this relationship or indicate none (add rows as needed)** | **Specifications/Comments (e.g., if payments were made to you or to your institution)** |
|---|---|---|---|
| | | **Time frame: Since the initial planning of the work** | |
| 1 | All support for the present manuscript (e.g., funding, provision of study materials, medical writing, article processing charges, etc.) **No time limit for this item.** | __X__ None | |
| | | **Time frame: past 36 months** | |
| 2 | Grants or contracts from any entity (if not indicated in item #1 above). | __X__ None | |
| 3 | Royalties or licenses | __X__ None | |

| | | | |
|---|---|---|---|
| 4 | Consulting fees | __X__ None | |
| 5 | Payment or honoraria for lectures, presentations, speakers bureaus, manuscript writing or educational events | __X__ None | |
| 6 | Payment for expert testimony | __X__ None | |
| 7 | Support for attending meetings and/or travel | __X__ None | |
| 8 | Patents planned, issued or pending | __X__ None | |
| 9 | Participation on a Data Safety Monitoring Board or Advisory Board | __X__ None | |
| 10 | Leadership or fiduciary role in other board, society, committee or advocacy group, paid or unpaid | __X__ None | |
| 11 | Stock or stock options | __X__ None | |
| 12 | Receipt of equipment, materials, drugs, medical writing, gifts or other services | __X__ None | |
| 13 | Other financial or non-financial interests | ____ None<br>Novartis Institutes for BioMedical Research | Currently employed at Novartis Institutes for BioMedical Research |

**Please place an "X" next to the following statement to indicate your agreement:**

**__X_ I certify that I have answered every question and have not altered the wording of any of the questions on this form.**

#### ICMJE DISCLOSURE FORM

**Date:__7/28/2021___________________________________________________________________________**
**Your Name:_____Joel Krier, MD_________________________________________________________________**
**Manuscript Title:_** Returning Actionable Genomic Results in a Research Biobank: Analytic Validity, Clinical Implementation and Resource Utilization
**_______________________________________________________________________________**
**Manuscript number (if known):_______________________________________________________________**

**In the interest of transparency, we ask you to disclose all relationships/activities/interests listed below that are related to the content of your manuscript. "Related" means any relation with for-profit or not-for-profit third parties whose interests may be affected by the content of the manuscript. Disclosure represents a commitment to transparency and does not necessarily indicate a bias. If you are in doubt about whether to list a relationship/activity/interest, it is preferable that you do so.**

**The following questions apply to the author's relationships/activities/interests as they relate to the current manuscript only.**

**The author's relationships/activities/interests should be defined broadly. For example, if your manuscript pertains to the epidemiology of hypertension, you should declare all relationships with manufacturers of antihypertensive medication, even if that medication is not mentioned in the manuscript.**

**In item #1 below, report all support for the work reported in this manuscript without time limit. For all other items, the time frame for disclosure is the past 36 months.**

| | | **Name all entities with whom you have this relationship or indicate none (add rows as needed)** | **Specifications/Comments (e.g., if payments were made to you or to your institution)** |
|---|---|---|---|
| | | **Time frame: Since the initial planning of the work** | |
| 1 | All support for the present manuscript (e.g., funding, provision of study materials, medical writing, article processing charges, etc.) **No time limit for this item.** | _X___ None | |
| | | **Time frame: past 36 months** | |
| 2 | Grants or contracts from any entity (if not indicated in item #1 above). | ____ Harvard Medical School | Clinical Genomics Analysis Platform |
| 3 | Royalties or licenses | X None | |

| | | | |
|---|---|---|---|
| 4 | Consulting fees | None<br>Gerson Lehrman Group | |
| 5 | Payment or honoraria for lectures, presentations, speakers bureaus, manuscript writing or educational events | X None | |
| 6 | Payment for expert testimony | X None | |
| 7 | Support for attending meetings and/or travel | None | GeneOne- travel to Brailizian College of Medical Genetics meeting, 2019 |
| 8 | Patents planned, issued or pending | X None | |
| 9 | Participation on a Data Safety Monitoring Board or Advisory Board | X None | |
| 10 | Leadership or fiduciary role in other board, society, committee or advocacy group, paid or unpaid | X None | |
| 11 | Stock or stock options | X None | |
| 12 | Receipt of equipment, materials, drugs, medical writing, gifts or other services | X None | |
| 13 | Other financial or non-financial interests | X None | |

**Please place an "X" next to the following statement to indicate your agreement:**

**X. I certify that I have answered every question and have not altered the wording of any of the questions on this form.**

**ICMJE DISCLOSURE FORM**

**Date:_07/29/21________________________________________________________________________________**
**Your Name: ___Matthew S Lebo________________________________________________________________________**
**Manuscript Title:_** Returning Actionable Genomic Results in a Research Biobank: Analytic Validity, Clinical Implementation and Resource Utilization
______________________________________________________________________________
**Manuscript number (if known):________________________________________________________________________**

**In the interest of transparency, we ask you to disclose all relationships/activities/interests listed below that are related to the content of your manuscript. "Related" means any relation with for-profit or not-for-profit third parties whose interests may be affected by the content of the manuscript. Disclosure represents a commitment to transparency and does not necessarily indicate a bias. If you are in doubt about whether to list a relationship/activity/interest, it is preferable that you do so.**

**The following questions apply to the author's relationships/activities/interests as they relate to the current manuscript only.**

**The author's relationships/activities/interests should be defined broadly. For example, if your manuscript pertains to the epidemiology of hypertension, you should declare all relationships with manufacturers of antihypertensive medication, even if that medication is not mentioned in the manuscript.**

**In item #1 below, report all support for the work reported in this manuscript without time limit. For all other items, the time frame for disclosure is the past 36 months.**

| | | **Name all entities with whom you have this relationship or indicate none (add rows as needed)** | **Specifications/Comments (e.g., if payments were made to you or to your institution)** |
|---|---|---|---|
| **Time frame: Since the initial planning of the work** | | | |
| 1 | All support for the present manuscript (e.g., funding, provision of study materials, medical writing, article processing charges, etc.) **No time limit for this item.** | ____ None | |
| | | NIH | Grants: HG008676 and HG008685 |
| **Time frame: past 36 months** | | | |
| 2 | Grants or contracts from any entity (if not indicated in item #1 above). | __X__ None | |
| 3 | Royalties or licenses | __X__ None | |

| | | | |
|---|---|---|---|
| 4 | Consulting fees | ___X_ None | |
| 5 | Payment or honoraria for lectures, presentations, speakers bureaus, manuscript writing or educational events | ____ None | |
| | | Association of Molecular Pathology | Honoraria for course design |
| 6 | Payment for expert testimony | __X__ None | |
| 7 | Support for attending meetings and/or travel | __X__ None | |
| 8 | Patents planned, issued or pending | __X__ None | |
| 9 | Participation on a Data Safety Monitoring Board or Advisory Board | ____ None | |
| | | NIH/NHGRI | SAB member for Human Pangenome Reference Consortium |
| 10 | Leadership or fiduciary role in other board, society, committee or advocacy group, paid or unpaid | ____ None | |
| | | Association of Molecular Pathology | Informatics Subdivision Leadership |
| 11 | Stock or stock options | __X__ None | |
| 12 | Receipt of equipment, materials, drugs, medical writing, gifts or other services | __X__ None | |
| 13 | Other financial or non-financial interests | __X__ None | |

**Please place an "X" next to the following statement to indicate your agreement:**

**__X_ I certify that I have answered every question and have not altered the wording of any of the questions on this form.**

**ICMJE DISCLOSURE FORM**

**Date:____07/28/2021__________________________________________________________________________**
**Your Name:_____Lisa M. Mahanta___________________________________________________________________**
**Manuscript Title:_** Returning Actionable Genomic Results in a Research Biobank: Analytic Validity, Clinical Implementation and Resource Utilization
**_____________________________________________________________________________**
**Manuscript number (if known):_________________________________________________________________**

**In the interest of transparency, we ask you to disclose all relationships/activities/interests listed below that are related to the content of your manuscript. "Related" means any relation with for-profit or not-for-profit third parties whose interests may be affected by the content of the manuscript. Disclosure represents a commitment to transparency and does not necessarily indicate a bias. If you are in doubt about whether to list a relationship/activity/interest, it is preferable that you do so.**

**The following questions apply to the author's relationships/activities/interests as they relate to the current manuscript only.**

**The author's relationships/activities/interests should be defined broadly. For example, if your manuscript pertains to the epidemiology of hypertension, you should declare all relationships with manufacturers of antihypertensive medication, even if that medication is not mentioned in the manuscript.**

**In item #1 below, report all support for the work reported in this manuscript without time limit. For all other items, the time frame for disclosure is the past 36 months.**

| | | **Name all entities with whom you have this relationship or indicate none (add rows as needed)** | **Specifications/Comments (e.g., if payments were made to you or to your institution)** |
|---|---|---|---|
| | | **Time frame: Since the initial planning of the work** | |
| 1 | All support for the present manuscript (e.g., funding, provision of study materials, medical writing, article processing charges, etc.) **No time limit for this item.** | ____ None<br>NIH HG008685 | eMERGE III NIH Grant funded part of this project |
| | | NIH HG008676 | eMERGE III Sequencing Core grant |
| | | **Time frame: past 36 months** | |
| 2 | Grants or contracts from any entity (if not indicated in item #1 above). | __X__ None | |
| 3 | Royalties or licenses | __X__ None | |

| | | |
|---|---|---|
| 4 | Consulting fees | _X___ None |
| 5 | Payment or honoraria for lectures, presentations, speakers bureaus, manuscript writing or educational events | _X___ None |
| 6 | Payment for expert testimony | _X___ None |
| 7 | Support for attending meetings and/or travel | __X__ None |
| 8 | Patents planned, issued or pending | _X___ None |
| 9 | Participation on a Data Safety Monitoring Board or Advisory Board | _X___ None |
| 10 | Leadership or fiduciary role in other board, society, committee or advocacy group, paid or unpaid | _X___ None |
| 11 | Stock or stock options | _X___ None |
| 12 | Receipt of equipment, materials, drugs, medical writing, gifts or other services | __X__ None |
| 13 | Other financial or non-financial interests | __X__ None |

**Please place an "X" next to the following statement to indicate your agreement:**

**_X__ I certify that I have answered every question and have not altered the wording of any of the questions on this form.**

**ICMJE DISCLOSURE FORM**

**Date:_July 27, 2021_______________________________________________________________________________**
**Your Name:_Marcie Steeves, MS, CGC__________________________________________________________________**
**Manuscript Title:_** Returning Actionable Genomic Results in a Research Biobank: Analytic Validity, Clinical Implementation and Resource Utilization
**__________________________________________________________________________________**
**Manuscript number (if known):____________________________________________________________________**

**In the interest of transparency, we ask you to disclose all relationships/activities/interests listed below that are related to the content of your manuscript. "Related" means any relation with for-profit or not-for-profit third parties whose interests may be affected by the content of the manuscript. Disclosure represents a commitment to transparency and does not necessarily indicate a bias. If you are in doubt about whether to list a relationship/activity/interest, it is preferable that you do so.**

**The following questions apply to the author's relationships/activities/interests as they relate to the current manuscript only.**

**The author's relationships/activities/interests should be defined broadly. For example, if your manuscript pertains to the epidemiology of hypertension, you should declare all relationships with manufacturers of antihypertensive medication, even if that medication is not mentioned in the manuscript.**

**In item #1 below, report all support for the work reported in this manuscript without time limit. For all other items, the time frame for disclosure is the past 36 months.**

| | | **Name all entities with whom you have this relationship or indicate none (add rows as needed)** | **Specifications/Comments (e.g., if payments were made to you or to your institution)** |
|---|---|---|---|
| | | **Time frame: Since the initial planning of the work** | |
| 1 | All support for the present manuscript (e.g., funding, provision of study materials, medical writing, article processing charges, etc.) **No time limit for this item.** | _X_ None | |
| | | **Time frame: past 36 months** | |
| 2 | Grants or contracts from any entity (if not indicated in item #1 above). | _X_ None | |
| 3 | Royalties or licenses | _X_ None | |

| | | |
|---|---|---|
| 4 | Consulting fees | _X_ None |
| 5 | Payment or honoraria for lectures, presentations, speakers bureaus, manuscript writing or educational events | _X_ None |
| 6 | Payment for expert testimony | _X_ None |
| 7 | Support for attending meetings and/or travel | _X_ None |
| 8 | Patents planned, issued or pending | _X_ None |
| 9 | Participation on a Data Safety Monitoring Board or Advisory Board | _X_ None |
| 10 | Leadership or fiduciary role in other board, society, committee or advocacy group, paid or unpaid | _X_ None |
| 11 | Stock or stock options | _X_ None |
| 12 | Receipt of equipment, materials, drugs, medical writing, gifts or other services | _X_ None |
| 13 | Other financial or non-financial interests | _X_ None |

**Please place an "X" next to the following statement to indicate your agreement:**

**_X_ I certify that I have answered every question and have not altered the wording of any of the questions on this Form.**

**ICMJE DISCLOSURE FORM**

**Date:___7/29/21________________________________________________________________________________**
**Your Name:___Nidhi Shah, MD______________________________________________________________________**
**Manuscript Title:_** Returning Actionable Genomic Results in a Research Biobank: Analytic Validity, Clinical Implementation and Resource Utilization
**______________________________________________________________________________**
**Manuscript number (if known):_____________________________________________________________________**

**In the interest of transparency, we ask you to disclose all relationships/activities/interests listed below that are related to the content of your manuscript. "Related" means any relation with for-profit or not-for-profit third parties whose interests may be affected by the content of the manuscript. Disclosure represents a commitment to transparency and does not necessarily indicate a bias. If you are in doubt about whether to list a relationship/activity/interest, it is preferable that you do so.**

**The following questions apply to the author's relationships/activities/interests as they relate to the current manuscript only.**

**The author's relationships/activities/interests should be defined broadly. For example, if your manuscript pertains to the epidemiology of hypertension, you should declare all relationships with manufacturers of antihypertensive medication, even if that medication is not mentioned in the manuscript.**

**In item #1 below, report all support for the work reported in this manuscript without time limit. For all other items, the time frame for disclosure is the past 36 months.**

| | | Name all entities with whom you have this relationship or indicate none (add rows as needed) | Specifications/Comments (e.g., if payments were made to you or to your institution) |
|---|---|---|---|
| | | **Time frame: Since the initial planning of the work** | |
| 1 | All support for the present manuscript (e.g., funding, provision of study materials, medical writing, article processing charges, etc.) **No time limit for this item.** | __√_ None | |
| | | **Time frame: past 36 months** | |
| 2 | Grants or contracts from any entity (if not indicated in item #1 above). | __√_ None | |
| 3 | Royalties or licenses | __√__ None | |

| | | |
|---|---|---|
| 4 | Consulting fees | __√__ None |
| 5 | Payment or honoraria for lectures, presentations, speakers bureaus, manuscript writing or educational events | __√__ None |
| 6 | Payment for expert testimony | __√__ None |
| 7 | Support for attending meetings and/or travel | __√__ None |
| 8 | Patents planned, issued or pending | __√__ None |
| 9 | Participation on a Data Safety Monitoring Board or Advisory Board | __√__ None |
| 10 | Leadership or fiduciary role in other board, society, committee or advocacy group, paid or unpaid | _√___ None |
| 11 | Stock or stock options | __√__ None |
| 12 | Receipt of equipment, materials, drugs, medical writing, gifts or other services | _√___ None |
| 13 | Other financial or non-financial interests | __√__ None |

**Please place an "X" next to the following statement to indicate your agreement:**

**_x__ I certify that I have answered every question and have not altered the wording of any of the questions on this form.**

### ICMJE DISCLOSURE FORM

**Date:__7/28/21_______________________________________________________________________**
**Your Name:_Robert Green_____________________________________________________________________**
**Manuscript Title:_** Returning Actionable Genomic Results in a Research Biobank: Analytic Validity, Clinical Implementation and Resource Utilization
**______________________________________________________________________________**
**Manuscript number (if known):_________________________________________________________________**

**In the interest of transparency, we ask you to disclose all relationships/activities/interests listed below that are related to the content of your manuscript. "Related" means any relation with for-profit or not-for-profit third parties whose interests may be affected by the content of the manuscript. Disclosure represents a commitment to transparency and does not necessarily indicate a bias. If you are in doubt about whether to list a relationship/activity/interest, it is preferable that you do so.**

**The following questions apply to the author's relationships/activities/interests as they relate to the current manuscript only.**

**The author's relationships/activities/interests should be defined broadly. For example, if your manuscript pertains to the epidemiology of hypertension, you should declare all relationships with manufacturers of antihypertensive medication, even if that medication is not mentioned in the manuscript.**

**In item #1 below, report all support for the work reported in this manuscript without time limit. For all other items, the time frame for disclosure is the past 36 months.**

| | | **Name all entities with whom you have this relationship or indicate none (add rows as needed)** | **Specifications/Comments (e.g., if payments were made to you or to your institution)** |
|---|---|---|---|
| **Time frame: Since the initial planning of the work** | | | |
| 1 | All support for the present manuscript (e.g., funding, provision of study materials, medical writing, article processing charges, etc.) **No time limit for this item.** | ____ None<br>NIH - HG008685 | eMERGE III NIH Grant funded part of this project |
| **Time frame: past 36 months** | | | |
| 2 | Grants or contracts from any entity (if not indicated in item #1 above). | ____ None<br>NIH | NIH-funded grants: R01 HL143295, R01 HG009922, RF1 AG047866, U19 HD077671 |
| 3 | Royalties or licenses | X None | |

| | | | |
|---|---|---|---|
| 4 | Consulting fees | _____ None | Dr. Green has received compensation for advising the following companies: AIA, Genomic Life, Grail, Humanity, Kneed Media, Plumcare, OptumLabs, Verily, VibrentHealth; and is co-founder of Genome Medical. |
| 5 | Payment or honoraria for lectures, presentations, speakers bureaus, manuscript writing or educational events | X None | |
| 6 | Payment for expert testimony | X None | |
| 7 | Support for attending meetings and/or travel | _____ None | Travel was reimbursed for meetings/events from the company Genome Medical. |
| 8 | Patents planned, issued or pending | X None | |
| 9 | Participation on a Data Safety Monitoring Board or Advisory Board | X None | |
| 10 | Leadership or fiduciary role in other board, society, committee or advocacy group, paid or unpaid | X None | |
| 11 | Stock or stock options | X None | |
| 12 | Receipt of equipment, materials, drugs, medical writing, gifts or other services | X None | |
| 13 | Other financial or non-financial interests | X None | |

**Please place an "X" next to the following statement to indicate your agreement:**

**X I certify that I have answered every question and have not altered the wording of any of the questions on this form.**

**ICMJE DISCLOSURE FORM**

**Date:____________7/27/21______________________________________________________________________**
**Your Name:____________Heidi Rehm___________________________________________________________________**
**Manuscript Title:_** Returning Actionable Genomic Results in a Research Biobank: Analytic Validity, Clinical Implementation and Resource Utilization
**_________________________________________________________________________**
**Manuscript number (if known):_______________________________________________________________**

**In the interest of transparency, we ask you to disclose all relationships/activities/interests listed below that are related to the content of your manuscript. "Related" means any relation with for-profit or not-for-profit third parties whose interests may be affected by the content of the manuscript. Disclosure represents a commitment to transparency and does not necessarily indicate a bias. If you are in doubt about whether to list a relationship/activity/interest, it is preferable that you do so.**

**The following questions apply to the author's relationships/activities/interests as they relate to the current manuscript only.**

**The author's relationships/activities/interests should be defined broadly. For example, if your manuscript pertains to the epidemiology of hypertension, you should declare all relationships with manufacturers of antihypertensive medication, even if that medication is not mentioned in the manuscript.**

**In item #1 below, report all support for the work reported in this manuscript without time limit. For all other items, the time frame for disclosure is the past 36 months.**

| | | **Name all entities with whom you have this relationship or indicate none (add rows as needed)** | **Specifications/Comments (e.g., if payments were made to you or to your institution)** |
|---|---|---|---|
| | | **Time frame: Since the initial planning of the work** | |
| 1 | All support for the present manuscript (e.g., funding, provision of study materials, medical writing, article processing charges, etc.) **No time limit for this item.** | _X__ None | |
| | | **Time frame: past 36 months** | |
| 2 | Grants or contracts from any entity (if not indicated in item #1 above). | _ X __ None | |
| 3 | Royalties or licenses | __ X __ None | |

| | | |
|---|---|---|
| 4 | Consulting fees | __ X __ None |
| 5 | Payment or honoraria for lectures, presentations, speakers bureaus, manuscript writing or educational events | _ X ___ None |
| 6 | Payment for expert testimony | __ X __ None |
| 7 | Support for attending meetings and/or travel | _ X ___ None |
| 8 | Patents planned, issued or pending | _ X ___ None |
| 9 | Participation on a Data Safety Monitoring Board or Advisory Board | _ X ___ None |
| 10 | Leadership or fiduciary role in other board, society, committee or advocacy group, paid or unpaid | _ X ___ None |
| 11 | Stock or stock options | _ X ___ None |
| 12 | Receipt of equipment, materials, drugs, medical writing, gifts or other services | __ X __ None |
| 13 | Other financial or non-financial interests | __ X __ None |

**Please place an "X" next to the following statement to indicate your agreement:**

_ X _ **I certify that I have answered every question and have not altered the wording of any of the questions on this form.**

#### ICMJE DISCLOSURE FORM

**Date:** July 29, 2021
**Your Name:** Janelle Shea
**Manuscript Title:**_ Returning Actionable Genomic Results in a Research Biobank: Analytic Validity, Clinical Implementation and Resource Utilization
______________________________________________________________________________
**Manuscript number (if known):**_________________________________________________________________

**In the interest of transparency, we ask you to disclose all relationships/activities/interests listed below that are related to the content of your manuscript. "Related" means any relation with for-profit or not-for-profit third parties whose interests may be affected by the content of the manuscript. Disclosure represents a commitment to transparency and does not necessarily indicate a bias. If you are in doubt about whether to list a relationship/activity/interest, it is preferable that you do so.**

**The following questions apply to the author's relationships/activities/interests as they relate to the current manuscript only.**

**The author's relationships/activities/interests should be defined broadly. For example, if your manuscript pertains to the epidemiology of hypertension, you should declare all relationships with manufacturers of antihypertensive medication, even if that medication is not mentioned in the manuscript.**

**In item #1 below, report all support for the work reported in this manuscript without time limit. For all other items, the time frame for disclosure is the past 36 months.**

| | | **Name all entities with whom you have this relationship or indicate none (add rows as needed)** | **Specifications/Comments (e.g., if payments were made to you or to your institution)** |
|---|---|---|---|
| | | **Time frame: Since the initial planning of the work** | |
| 1 | All support for the present manuscript (e.g., funding, provision of study materials, medical writing, article processing charges, etc.) **No time limit for this item.** | __X__ None | |
| | | **Time frame: past 36 months** | |
| 2 | Grants or contracts from any entity (if not indicated in item #1 above). | _X___ None | |
| 3 | Royalties or licenses | _X___ None | |

| | | |
|---|---|---|
| 4 | Consulting fees | _X___ None |
| 5 | Payment or honoraria for lectures, presentations, speakers bureaus, manuscript writing or educational events | __X__ None |
| 6 | Payment for expert testimony | _X___ None |
| 7 | Support for attending meetings and/or travel | __X__ None |
| 8 | Patents planned, issued or pending | __X__ None |
| 9 | Participation on a Data Safety Monitoring Board or Advisory Board | __X__ None |
| 10 | Leadership or fiduciary role in other board, society, committee or advocacy group, paid or unpaid | __X__ None |
| 11 | Stock or stock options | _X___ None |
| 12 | Receipt of equipment, materials, drugs, medical writing, gifts or other services | _X___ None |
| 13 | Other financial or non-financial interests | _X___ None |

**Please place an "X" next to the following statement to indicate your agreement:**

**_X I certify that I have answered every question and have not altered the wording of any of the questions on this form.**

**ICMJE DISCLOSURE FORM**

**Date:____07/30/21**
**Your Name:__ Jordan W. Smoller, MD, ScD__________**
**Manuscript Title:_** Returning Actionable Genomic Results in a Research Biobank: Analytic Validity, Clinical Implementation and Resource Utilization
**_______________________________________________________________________________**
**Manuscript number (if known):_______________________________________________________________________**

**In the interest of transparency, we ask you to disclose all relationships/activities/interests listed below that are related to the content of your manuscript. "Related" means any relation with for-profit or not-for-profit third parties whose interests may be affected by the content of the manuscript. Disclosure represents a commitment to transparency and does not necessarily indicate a bias. If you are in doubt about whether to list a relationship/activity/interest, it is preferable that you do so.**

**The following questions apply to the author's relationships/activities/interests as they relate to the current manuscript only.**

**The author's relationships/activities/interests should be defined broadly. For example, if your manuscript pertains to the epidemiology of hypertension, you should declare all relationships with manufacturers of antihypertensive medication, even if that medication is not mentioned in the manuscript.**

**In item #1 below, report all support for the work reported in this manuscript without time limit. For all other items, the time frame for disclosure is the past 36 months.**

| | | **Name all entities with whom you have this relationship or indicate none (add rows as needed)** | **Specifications/Comments (e.g., if payments were made to you or to your institution)** |
|---|---|---|---|
| | | **Time frame: Since the initial planning of the work** | |
| 1 | All support for the present manuscript (e.g., funding, provision of study materials, medical writing, article processing charges, etc.) **No time limit for this item.** | ____ None<br>NIH HG008685 | eMERGE III NIH Grant funded part of this project |
| | | **Time frame: past 36 months** | |
| 2 | Grants or contracts from any entity (if not indicated in item #1 above). | __X__ None | |
| 3 | Royalties or licenses | __X__ None | |

| | | | |
|---|---|---|---|
| 4 | Consulting fees | ___X_ None | |
| 5 | Payment or honoraria for lectures, presentations, speakers bureaus, manuscript writing or educational events | __X__ None | |
| 6 | Payment for expert testimony | __X__ None | |
| 7 | Support for attending meetings and/or travel | __X__ None | |
| 8 | Patents planned, issued or pending | __X__ None | |
| 9 | Participation on a Data Safety Monitoring Board or Advisory Board | ____ NHGRI | Member of DSMB for the Implementing Genomics in Practice (IGNITE II) program |
| 10 | Leadership or fiduciary role in other board, society, committee or advocacy group, paid or unpaid | __X__ None | |
| 11 | Stock or stock options | __X__ None | |
| 12 | Receipt of equipment, materials, drugs, medical writing, gifts or other services | __X__ None | |
| 13 | Other financial or non-financial interests | ____ 23andme | PI of a collaborative study of the genetics of depression and bipolar disorder sponsored by 23andMe for which 23andMe provides analysis time as in-kind support but no payments. |

**Please place an "X" next to the following statement to indicate your agreement:**

**_X__ I certify that I have answered every question and have not altered the wording of any of the questions on this form.**

#### ICMJE DISCLOSURE FORM

**Date:_7/29/2021_______________________________________________________________________________**
**Your Name:__Melissa K. Uveges____________________________________________________________________________**
**Manuscript Title:_** Returning Actionable Genomic Results in a Research Biobank: Analytic Validity, Clinical Implementation and Resource Utilization
**_____________________________________________________________________________**
**Manuscript number (if known):___________________________________________________________________**

**In the interest of transparency, we ask you to disclose all relationships/activities/interests listed below that are related to the content of your manuscript. "Related" means any relation with for-profit or not-for-profit third parties whose interests may be affected by the content of the manuscript. Disclosure represents a commitment to transparency and does not necessarily indicate a bias. If you are in doubt about whether to list a relationship/activity/interest, it is preferable that you do so.**

**The following questions apply to the author's relationships/activities/interests as they relate to the current manuscript only.**

**The author's relationships/activities/interests should be defined broadly. For example, if your manuscript pertains to the epidemiology of hypertension, you should declare all relationships with manufacturers of antihypertensive medication, even if that medication is not mentioned in the manuscript.**

**In item #1 below, report all support for the work reported in this manuscript without time limit. For all other items, the time frame for disclosure is the past 36 months.**

| | | **Name all entities with whom you have this relationship or indicate none (add rows as needed)** | **Specifications/Comments (e.g., if payments were made to you or to your institution)** |
|---|---|---|---|
| | | **Time frame: Since the initial planning of the work** | |
| 1 | All support for the present manuscript (e.g., funding, provision of study materials, medical writing, article processing charges, etc.) **No time limit for this item.** | _X___ None | |
| | | **Time frame: past 36 months** | |
| 2 | Grants or contracts from any entity (if not indicated in item #1 above). | __X__ None | |
| 3 | Royalties or licenses | __X__ None | |

| | | |
|---|---|---|
| 4 | Consulting fees | _X___ None |
| 5 | Payment or honoraria for lectures, presentations, speakers bureaus, manuscript writing or educational events | _X___ None |
| 6 | Payment for expert testimony | _X___ None |
| 7 | Support for attending meetings and/or travel | __X__ None |
| 8 | Patents planned, issued or pending | __X__ None |
| 9 | Participation on a Data Safety Monitoring Board or Advisory Board | _X___ None |
| 10 | Leadership or fiduciary role in other board, society, committee or advocacy group, paid or unpaid | _X___ None |
| 11 | Stock or stock options | _X___ None |
| 12 | Receipt of equipment, materials, drugs, medical writing, gifts or other services | __X__ None |
| 13 | Other financial or non-financial interests | __X__ None |

**Please place an "X" next to the following statement to indicate your agreement:**

**_X__ I certify that I have answered every question and have not altered the wording of any of the questions on this form.**

#### ICMJE DISCLOSURE FORM

**Date:____07/29/2021______________________________________________________________________________**

**Your Name:__Scott T. Weiss_________________________________________________________________________**
**Manuscript Title:_** Returning Actionable Genomic Results in a Research Biobank: Analytic Validity, Clinical Implementation and Resource Utilization

**______________________________________________________________________________**
**Manuscript number (if known):____________________________________________________________________**

**In the interest of transparency, we ask you to disclose all relationships/activities/interests listed below that are related to the content of your manuscript. "Related" means any relation with for-profit or not-for-profit third parties whose interests may be affected by the content of the manuscript. Disclosure represents a commitment to transparency and does not necessarily indicate a bias. If you are in doubt about whether to list a relationship/activity/interest, it is preferable that you do so.**

**The following questions apply to the author's relationships/activities/interests as they relate to the current manuscript only.**

**The author's relationships/activities/interests should be defined broadly. For example, if your manuscript pertains to the epidemiology of hypertension, you should declare all relationships with manufacturers of antihypertensive medication, even if that medication is not mentioned in the manuscript.**

**In item #1 below, report all support for the work reported in this manuscript without time limit. For all other items, the time frame for disclosure is the past 36 months.**

| | | **Name all entities with whom you have this relationship or indicate none (add rows as needed)** | **Specifications/Comments (e.g., if payments were made to you or to your institution)** |
|---|---|---|---|
| | | **Time frame: Since the initial planning of the work** | |
| 1 | All support for the present manuscript (e.g., funding, provision of study materials, medical writing, article processing charges, etc.) **No time limit for this item.** | ____ None<br>NIH - HG008685 | |
| | | **Time frame: past 36 months** | |
| 2 | Grants or contracts from any entity (if not indicated in item #1 above). | ____ None<br>NIH - HG008685 | |

| | | |
|---|---|---|
| 3 | Royalties or licenses | __X__ None |
| 4 | Consulting fees | __X__ None |
| 5 | Payment or honoraria for lectures, presentations, speakers bureaus, manuscript writing or educational events | ____ None<br>I have received payment from UpToDate |
| 6 | Payment for expert testimony | __X__ None |
| 7 | Support for attending meetings and/or travel | ___X_ None |
| 8 | Patents planned, issued or pending | __X__ None |
| 9 | Participation on a Data Safety Monitoring Board or Advisory Board | __X__ None |
| 10 | Leadership or fiduciary role in other board, society, committee or advocacy group, paid or unpaid | __X__ None |
| 11 | Stock or stock options | __X__ None |
| 12 | Receipt of equipment, materials, drugs, medical writing, gifts or other services | __X__ None |
| 13 | Other financial or non-financial interests | __X__ None |

**Please place an "X" next to the following statement to indicate your agreement:**

**_X__ I certify that I have answered every question and have not altered the wording of any of the questions on this form.**
